# Elevated Atherogenicity in Long COVID: A Systematic Review and Meta-Analysis

**DOI:** 10.1101/2025.01.06.25320068

**Authors:** Abbas F. Almulla, Yanin Thipakorn, Yingqian Zhang, Michael Maes

## Abstract

**Background:** Long COVID (LC) is a complex, multi-organ syndrome that persists following recovery from the acute phase of coronavirus infection. Cardiovascular involvement is frequently reported in LC, often accompanied by a spectrum of related symptoms. Dysregulated lipid profiles and elevated atherogenic indices have been implicated in LC, yet no comprehensive systematic review and meta-analysis has specifically addressed these biomarkers.

**Objective:** This study aims to systematically evaluate atherogenic indices and lipid-related biomarkers in individuals with LC compared to healthy controls.

**Methods:** A systematic search was conducted in databases including PubMed, Google Scholar, SCOPUS, and SciFinder from September to November 2024. Eligible studies reported lipid biomarker data for LC patients and controls, yielding 44 studies encompassing 8,114 participants (3,353 LC patients and 4,761 controls).

**Results:** LC patients exhibited significant elevations in Castelli Risk Indexes 1 (standardized mean difference, SMD = 0.199; 95% confidence intervals, CI: 0.087–0.312) and 2 (SMD = 0.202; 95% CI: 0.087–0.318). Atherogenic ratios, including triglyceride (TG)/high-density lipoprotein (HDL) (SMD = 0.294; 95% CI: 0.155–0.433), (TG + low-density lipoprotein, LDL + very low-density lipoprotein, VLDL)/(HDL + apolipoprotein, ApoA) (SMD = 0.264; 95% CI: 0.145–0.383), and ApoB/ApoA (SMD = 0.515; 95% CI: 0.233–0.796), were also significantly elevated. Additionally, LC patients demonstrated increased levels of LDL, total cholesterol, triglycerides, and ApoB, alongside reduced HDL and ApoA levels. Results were free from publication bias.

**Conclusion:** LC is associated with a pro-atherogenic lipid profile, marked by increased atherogenic components and decreased protective lipid biomarkers. These findings highlight a potential heightened risk for cardiovascular complications in LC patients, warranting further clinical and mechanistic investigations.

## Introduction

The comprehension of post-viral syndromes has advanced notably after the Severe acute respiratory syndrome coronavirus 2 (SARS-CoV-2) pandemic, leading to the emergence of Long Coronavirus (LC) disease, a multifaceted and multisystem condition impacting around 10–30% of patients who have recovered from the acute infection g(Nalbandian, Sehgal et al. 2021, Qin, Zhang et al. 2024). LC presents significant challenges to healthcare systems and negatively impacts patients’ quality of life (Maes, Al-Rubaye et al. 2022, Katz, Bach et al. 2023) across multiple organ systems, including cardiovascular, respiratory, neurological, gastrointestinal, renal, musculoskeletal, endocrine, and immune systems (Li, Zhou et al. 2023). Despite progress in Coronavirus (COVID-19) research, the mechanisms through which it impacts systemic organs after recovery, especially the cardiovascular system, are not well understood, underscoring the need for focused investigation.

Key features of LC include neuropsychiatric symptoms, such as affective symptoms, fatigue and memory impairment, as well as systemic dysfunctions, including metabolic abnormalities (Dennis, Wamil et al. 2021, Lopez-Leon, Wegman-Ostrosky et al. 2021, Mehandru and Merad 2022). Several biological disturbances and pathways were discovered and believed to contribute the pathophysiology of LC disease including but not limited to upregulated systemic immune-inflammatory response (Almulla, Al-Hakeim et al. 2023, Almulla, Thipakorn et al. 2024a), elevated oxidative and nitrosative stress, insulin resistance (Al-Hakeim, Al-Rubaye et al. 2023, Maes, Almulla et al. 2023), gut dysbiosis (Su, Lau et al. 2024), mitochondrial dysfunction (Molnar, Lehoczki et al. 2024), which are all linked to a pro-atherogenic milieu (Al-Hakeim, Al-Rubaye et al. 2023). Other contributors include neuroinflammation and autoimmunity targeting central nervous system proteins (Almulla, Maes et al. 2024d), lowered tryptophan and activated kynurenine pathway (Almulla, Thipakorn et al. 2024b), and reactivation of latent viruses like human herpesvirus 6 (HHV-6) and Epstein–Barr virus (EBV) (Vojdani, Almulla et al. 2024). Nonetheless, the precise mechanisms through which these processes interact to elevate cardiovascular risk, especially via endothelial dysfunction and atherogenesis, are not yet thoroughly investigated.

Hepatocellular damage is another prominent feature of LC that may contribute to these processes. We recently published that LC is associated with significant liver damage marked by biomarkers such as transaminases enzymes and acute phase reactants (Almulla, Thipakorn et al. 2024c). The liver plays a central role in lipid homeostasis and metabolic regulation and its damage in LC could significantly disrupts reverse cholesterol transport (RCT) mechanisms mediated by high-density lipoprotein (HDL), apolipoprotein A (ApoA), paraoxonase 1 (PON1), and lecithin:cholesterol acyltransferase (LCAT), contributing to cholesterol accumulation, activating immune-inflammatory and oxidative stress pathways thereby increasing cardiovascular risk (Fisher, Feig et al. 2012, Almulla, Thipakorn et al. 2023, Almulla, Thipakorn et al. 2024c). Systemic inflammation further aggravates atherogenicity by impairing hepatic lipoprotein synthesis and promoting atherogenic lipid changes, such as increased triglycerides (TGs) and reduced HDL cholesterol (McGillicuddy, de la Llera Moya et al. 2009, Stuķ 2009, Jaganathan, Ravindran et al. 2018). These findings underscore the complex interplay between immune-inflammatory processes, liver injury, and lipid dysregulation in LC, further implicating these pathways in cardiovascular risk. However, the precise mechanisms through which systemic inflammation and immune dysregulation influence lipid metabolism to promote atherogenesis in LC remain inadequately investigated.

The cardiovascular risks following acute stage of COVID disease are substantial, as studies indicate a rise in cerebrovascular disorders, arrhythmias, heart disease, and thromboembolic events among survivors of COVID-19 (Xie, Xu et al. 2022). Dyslipidemia has emerged as a key mediator of these risks, characterized by abnormal lipid profiles, including but not limited to elevated total cholesterol, low-density lipoprotein (LDL), and TGs, alongside decreased HDL (Menezes, Lima et al. 2023, Xu, Xie et al. 2023, Tsampasian, Bäck et al. 2024). Additional lipid-related abnormalities include elevated apolipoprotein (Apo)B and reduced levels of ApoA and LCAT. These lipid abnormalities promote plaque formation, vascular inflammation, and ischemic injury, exacerbating cardiovascular complications (Bhargava, de la Puente-Secades et al. 2022). In addition, advanced lipidomic analyses have revealed significant alterations in phospholipids and sphingolipids in LC patients, further suggesting that immune-metabolic crosstalk is a key driver of LC-associated dyslipidemia (Garrido et al., 2024). Unlike traditional lipid markers, advanced atherogenic indices, such as Castelli’s Risk Index (CRI1 and CRI2), TG/HDL, and ApoB/ApoA ratios, offer a deeper understanding of the balance between atherogenic and anti-atherogenic lipid profiles (Jirakran et al., 2024). These indices are underexplored in LC, highlighting a significant gap in the understanding of cardiovascular risks related to this condition.

Nevertheless, no previous meta-analysis has systematically evaluated lipid biomarkers and atherogenic indices in LC. Hence, this study seeks to examine atherogenic indices, including CRI1, CRI2, TG/HDL, (TG+LDL+VLDL)/(HDL+ApoA), and ApoB/ApoA ratios, alongside individual lipid biomarkers, including HDL, LDL, VLDL, total cholesterol, TG, ApoA, and ApoB in LC patients compared to healthy controls. By focusing on these indices, the study aims to discuss the mechanisms underlying lipid dysregulation and its association with immune-inflammatory responses in LC, elucidating the elevated risk of cardiovascular complications. This research may contribute to targeted lipid-modifying therapies, potentially improving patient outcomes.

## Materials and method

This study adhered to rigorous methodological standards, incorporating the PRISMA 2020 guidelines, the Cochrane Handbook for Systematic Reviews of Interventions, and the MOOSE guidelines for observational studies. The primary analyses compared LC patients with normal controls (NC), focusing on atherogenic indices such as CRI1, CRI2, as well as ratios including TG/HDL, (TG+LDL+VLDL)/(HDL+ApoA), and ApoB/ApoA. The secondary analyses comprise individual lipid biomarkers, including HDL, TCHO, TGs, LDL, VLDL, ApoB, and ApoA, were thoroughly evaluated.

## Search strategy

A systematic search was conducted across electronic databases, including PubMed/MEDLINE, Google Scholar, and SciFinder, to gather comprehensive data on atherogenic indices and lipid biomarkers in LC. The search was conducted from September 10 to the end of November 2024, employing predefined keywords and MeSH terms as specified in ESF, Table 1. We examined reference lists from relevant studies and prior systematic reviews if available to identify additional significant publications, ensuring comprehensive coverage.

**Table 1.**
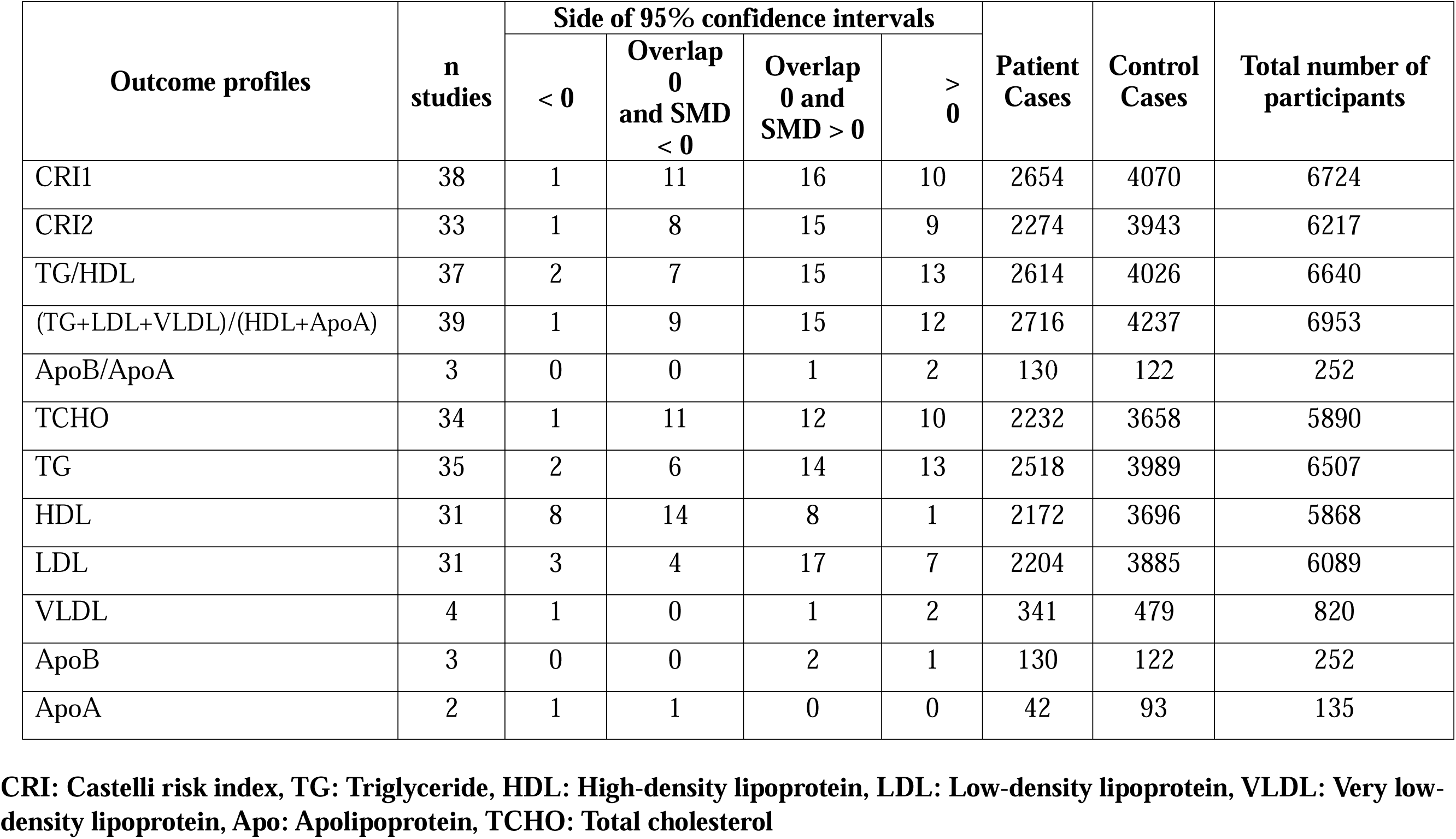
The outcomes and number of patients with Long COVID (LC) and normal controls along with the side of standardized mean difference (SMD) and the 95% confidence intervals with respect to zero SMD.

## Eligibility Criteria

The eligibility criteria included a comprehensive search for pertinent studies, emphasizing peer-reviewed publications in English. To ensure comprehensive data inclusion, grey literature and studies published in Thai, French, Spanish, Turkish, Hungarian, German, Ukrainian, Italian, Russian, and Arabic were considered. Studies were deemed eligible when they satisfied the following criteria: they utilized an observational design (case-control or cohort) with suitable control groups, examined atherogenic indices and individual lipid biomarkers in liver cancer patients diagnosed according to WHO criteria (World Health Organization 2021), and incorporated baseline biomarker evaluations followed by longitudinal follow-up. Exclusion criteria comprised studies employing unconventional biological samples, including hair, saliva, platelet-rich plasma, or cerebrospinal fluid (CSF); studies centered on genetic or translational research; and studies that did not include a control group. Furthermore, studies lacking data on the mean and standard deviation (SD) or standard error (SE) of biomarkers were excluded unless the required data could be acquired from the authors upon request or derived using established methods and tools, such as WebPlotDigitizer or other resources (e.g., hkbu.edu.hk).

## Primary and secondary outcomes

This meta-analysis highlighted several key indices and ratio regarding atherogenicity, including CRI1, CRI2, TG/HDL, (TG+LDL+VLDL)/(HDL+ApoA), and ApoB/ApoA ratios, as summarized in **Table 1**. In addition to these composite indices, individual biomarkers encompassing HDL, TCHO, TGs, LDL, VLDL, ApoB, and ApoA were analyzed as secondary outcomes.

## Screening and Data Extraction

To initiate this meta-analysis, two researchers, AA and YT, screened study titles and abstracts against predefined inclusion criteria. Full texts of potentially relevant studies were obtained for detailed evaluation, while those that did not meet exclusion criteria were excluded. Data extraction was systematically performed using a custom-designed Excel sheet. Information recorded included study metadata (author names, publication dates), lipid-related biomarker values (means and standard deviations), participant numbers in LC patients and control groups, and overall sample. The dataset also captured additional variables such as study design, biological sample types (e.g., serum, plasma, cerebrospinal fluid, blood cells, brain tissue), durations following post-COVID infection, intensive care unit (ICU) admission during acute infection, age, gender distribution, and study location and latitude. Any disagreements during data entry or interpretation were resolved by consulting the senior author, MM.

The methodological quality of the studies was assessed using the Immunological Confounder Scale (ICS), originally introduced by (Andrés-Rodríguez, Borràs et al. 2020) and later customized by MM to evaluate atherogenicity in LC. The ICS consists of two evaluation tools: the Quality Scale and the Redpoints Scale, detailed in Table 2 of ESF-1. These tools have been successfully employed in prior meta-analyses focused on immune activation, kynurenine pathway, and liver dysfunction biomarkers in LC (Almulla, Thipakorn et al. 2024a, Almulla,

**Table 2.** Results of meta-analysis performed on several outcomes (atherogenic indices and lipid profile) variables with combined different media and separately.

| Outcome feature sets | n | Groups | SMD | 95% CI | z | p | Q | df | p | I<sup>2</sup> (%) | $\tau^2$ | T |
| --- | --- | --- | --- | --- | --- | --- | --- | --- | --- | --- | --- | --- |
| CRI1 | 38 | Overall | 0.199 | 0.087;0.312 | 3.476 | 0.001 | 130.710 | 37 | 0.000 | 71.693 | 0.078 | 0.280 |
| CRI2 | 33 | Overall | 0.202 | 0.087;0.318 | 3.432 | 0.001 | 108.774 | 32 | 0.000 | 70.581 | 0.071 | 0.266 |
| TG/HDL | 37 | Overall | 0.294 | 0.155;0.433 | 4.139 | 0.000 | 194.141 | 36 | 0.000 | 81.457 | 0.136 | 0.369 |
| (TG+LDL+VLDL)/(HDL+ApoA) | 39 | Overall | 0.264 | 0.145;0.383 | 4.358 | 0.000 | 157.836 | 38 | 0.000 | 75.924 | 0.097 | 0.311 |
| ApoB/ApoA | 3 | Overall | 0.515 | 0.233;0.796 | 3.585 | 0.000 | 0.328 | 2 | 0.849 | 0.000 | 0.000 | 0.000 |
| HDL | 31 | Overall | -0.236 | -0.386; -0.087 | -3.093 | 0.002 | 162.057 | 30 | 0.000 | 81.488 | 0.131 | 0.361 |
| TG | 35 | Overall | 0.349 | 0.151;0.574 | 3.460 | 0.001 | 375.450 | 34 | 0.000 | 90.944 | 0.304 | 0.551 |
| TCHO | 34 | Overall | 0.156 | 0.036;0.276 | 2.545 | 0.011 | 112.036 | 33 | 0.000 | 70.545 | 0.079 | 0.280 |
| LDL | 31 | Overall | 0.196 | 0.070;0.323 | 3.048 | 0.002 | 119.642 | 30 | 0.000 | 74.925 | 0.085 | 0.291 |
| VLDL | 4 | Overall | 0.409 | -0.408;1.225 | 0.981 | 0.327 | 89.919 | 3 | 0.000 | 96.664 | 0.669 | 0.818 |
| ApoB | 3 | Overall | 0.301 | 0.023;0.580 | 2.119 | 0.034 | 1.183 | 3 | 0.553 | 0.000 | 0.000 | 0.000 |
| ApoA | 2 | Overall | -0.749 | -1.129;-0.370 | -3.874 | 0.000 | 0.000 | 1 | 0.985 | 0.000 | 0 | 0 |

**CRI:** Castelli risk index, **TG:** Triglyceride, **HDL:** High-density lipoprotein, **LDL:** Low-density lipoprotein, **VLDL:** Very low-density lipoprotein, **Apo:** Apolipoprotein, **TCHO:** Total cholesterol

Thipakorn et al. 2024b, Almulla, Thipakorn et al. 2024c), as well as lipid peroxidation and tryptophan metabolism in affective disorders (Almulla, Thipakorn et al. 2022, Almulla, Thipakorn et al. 2023). The Quality Scale assesses factors including sample size, control of confounders, sampling methods, providing scores ranging from 0 (lowest quality) to 10 (highest quality). The Redpoints Scale evaluates biases in immune activation studies in LC, with scores from 0 (ideal control) to 26 (no control).

## Data Analysis

The meta-analysis utilized CMA V4 software and followed the PRISMA guidelines. The requirements for inclusion, as specified in Table 3 of the ESF, mandated a minimum of two studies for each atherogenic indicator and lipid biomarker. For the calculation of the CRI, CRI2 and TG/HDL indices, complete dependence was assumed. Effect sizes were designated as positive for total cholesterol, TG or LDL and negative for HDL. Similarly, the (TG+LDL+VLDL)/(HDL+ApoA) ratio was calculated with positive effect sizes assigned to TG, LDL, and VLDL, and negative effect sizes to HDL and ApoA. Effect sizes for ApoB were defined as positive, while those for ApoA were negative.

**Table 3.**
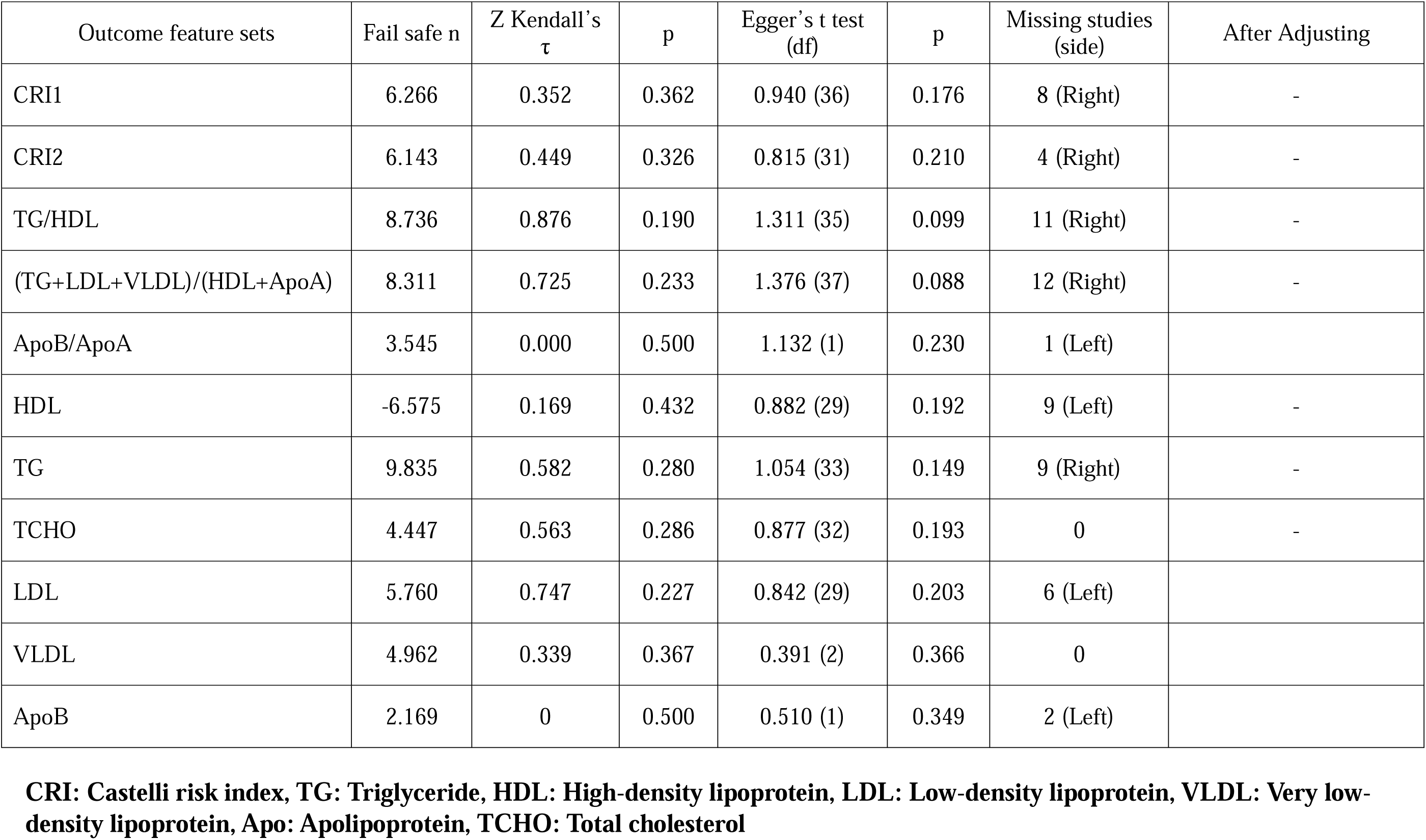
Results on publication bias.

| Outcome feature sets | Fail safe n | Z Kendall's $\tau$ | p | Egger's t test (df) | p | Missing studies (side) | After Adjusting |
| --- | --- | --- | --- | --- | --- | --- | --- |
| CRI1 | 6.266 | 0.352 | 0.362 | 0.940 (36) | 0.176 | 8 (Right) | - |
| CRI2 | 6.143 | 0.449 | 0.326 | 0.815 (31) | 0.210 | 4 (Right) | - |
| TG/HDL | 8.736 | 0.876 | 0.190 | 1.311 (35) | 0.099 | 11 (Right) | - |
| (TG+LDL+VLDL)/(HDL+ApoA) | 8.311 | 0.725 | 0.233 | 1.376 (37) | 0.088 | 12 (Right) | - |
| ApoB/ApoA | 3.545 | 0.000 | 0.500 | 1.132 (1) | 0.230 | 1 (Left) | - |
| HDL | -6.575 | 0.169 | 0.432 | 0.882 (29) | 0.192 | 9 (Left) | - |
| TG | 9.835 | 0.582 | 0.280 | 1.054 (33) | 0.149 | 9 (Right) | - |
| TCHO | 4.447 | 0.563 | 0.286 | 0.877 (32) | 0.193 | 0 | - |
| LDL | 5.760 | 0.747 | 0.227 | 0.842 (29) | 0.203 | 6 (Left) | - |
| VLDL | 4.962 | 0.339 | 0.367 | 0.391 (2) | 0.366 | 0 | - |
| ApoB | 2.169 | 0 | 0.500 | 0.510 (1) | 0.349 | 2 (Left) | - |

**CRI:** Castelli risk index, **TG:** Triglyceride, **HDL:** High-density lipoprotein, **LDL:** Low-density lipoprotein, **VLDL:** Very low-density lipoprotein, **Apo:** Apolipoprotein, **TCHO:** Total cholesterol

A random-effects model, based on constrained maximum likelihood, was employed to pool effect sizes, with statistical significance defined by a p-value of less than 0.05. Effect sizes were reported as standardized mean differences (SMD) with 95% confidence intervals (CIs). Following Cohen’s classification (Cohen, 2013), SMDs of 0.80, 0.50, and 0.20 indicated large, moderate, and small effect sizes, respectively. Heterogeneity was evaluated using tau-squared as the primary measure, with Q and I² values used as secondary indicators. Meta-regression analyses were conducted to identify potential sources of heterogeneity.

To ensure robustness of the findings, leave-one-out sensitivity analyses were performed. Publication bias was assessed using the fail-safe N approach, continuity-corrected Kendall’s tau, and Egger’s regression intercept, with the latter two methods providing one-tailed p-values. The trim-and-fill method was applied to address asymmetry detected by Egger’s test, and adjusted effect sizes were computed for missing studies. Funnel plots, incorporating both observed and imputed studies, were used to visualize small-study effects.

## Results

### Study outcomes

This study involved a systematic search utilizing targeted keywords and MeSH terms across various databases, including PubMed, Google Scholar, SciFinder, and SCOPUS (refer to ESF Table 1). The initial search yielded 2433 articles, as outlined in the PRISMA flow chart (**Figure 1**). After a meticulous screening process to eliminate irrelevant and duplicate entries, 102 studies remained. Of these, 44 studies met the predefined inclusion criteria and were subsequently included in the meta-analysis, adhering to the specified guidelines (Gameil, Marzouk et al. 2021, Holmes, Wist et al., Ломайчиков et al. 2021, Alfadda, Rafiullah et al. 2022, Aparisi, Martín-Fernández et al. 2022, Chudzik, Lewek et al. 2022, de Oliveira, de Ávila et al. 2022, Duran, Kurtipek et al. 2022, Labarca, Henríquez-Beltrán et al. 2022, Sumbalová, Kucharská et al. 2022, Tong, Yan et al. 2022, Verma, Ramayya et al. 2022, Abdulaziz Alsufyani 2023, Alshehri, AlQahtani et al. 2023, Berezhnoy, Bissinger et al. 2023, Erol, Tezcan et al. 2023, Kalinskaya, Vorobyeva et al. 2023, Kovarik, Bileck et al. 2023, Mora, Kogut et al. 2023, Mouchati, Durieux et al. 2023, Oikonomou, Lampsas et al. 2023, Paris, Palomba et al. 2023, Paul, Paul et al. 2023, Silva, Pereira et al. 2023, Szczerbiń Okruszko et al. 2023, Tudoran, Bende et al. 2023, Vollrath, Matits et al. 2023, Vyas, Joshi et al. 2023, Xuereb, Borg et al. 2023, Yamamoto, Otsuka et al. 2023, Zisis, Durieux et al. 2023, Agafonova, Elovikova et al. 2024, Al-Zadjali, Al-Lawati et al. 2024, Davico, Martín et al. 2024, Emiroglu, Dicle et al. 2024, Garrido, Castillo-Peinado et al. 2024, Grote, Schaefer et al. 2024, Korkmaz, Çınar et al. 2024, Kuryłowicz, Babicki et al. 2024, Liu and Kang 2024, Oliván-Blázquez, Bona-Otal et al. 2024, Santana-de Anda, Torres-Ruiz et al. 2024, Stepanova, Driianska et al. 2024, Zerón-Rugerio, Zaragozá et al. 2024).

**Figure 1.**
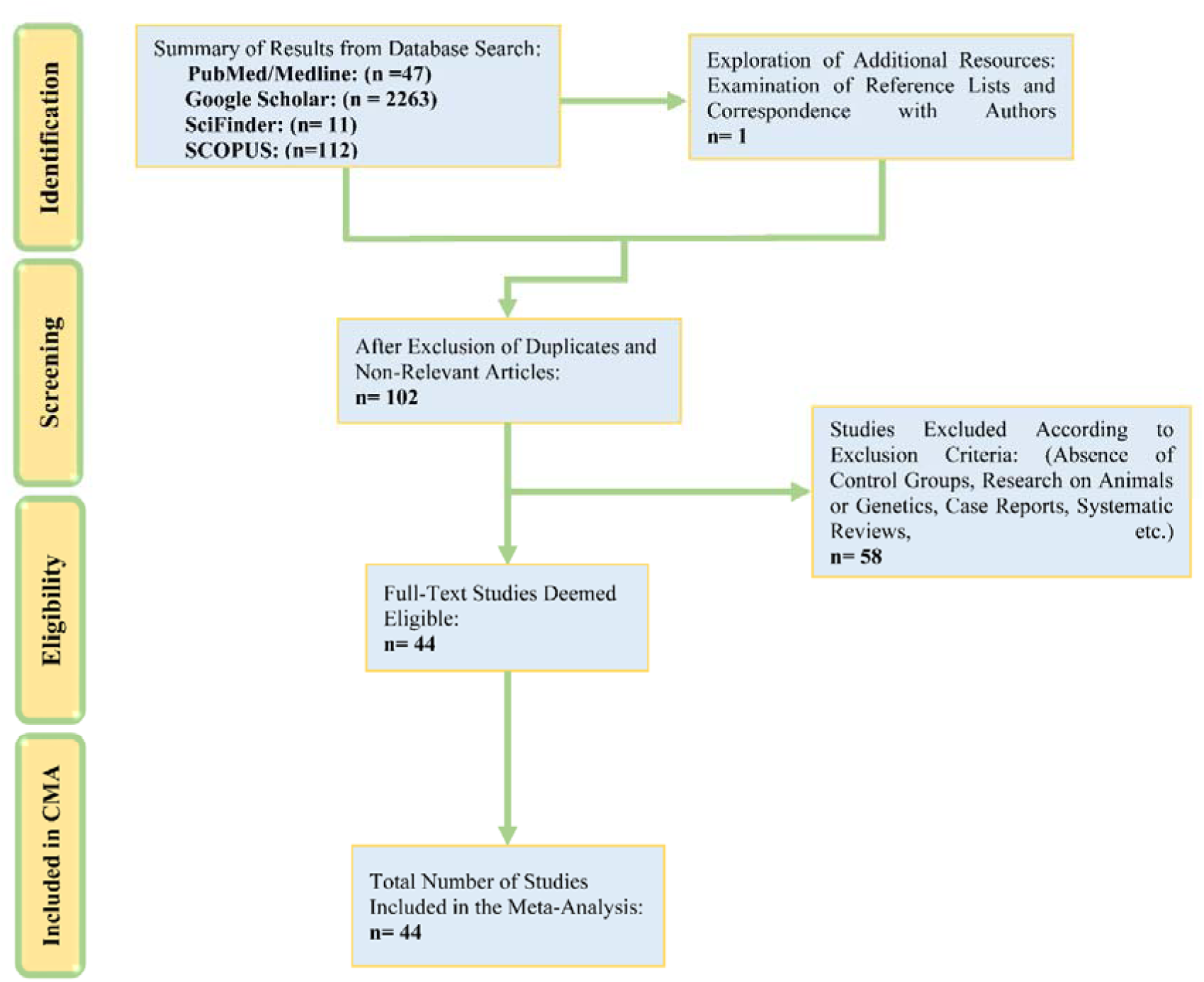
The PRISMA flow chart.

The meta-analysis incorporated data from 8114 participants, consisting of 3353 individuals with LC disease and 4761 NC. Participant ages ranged from 11 to 71.5 years. Most of the studies utilized routine laboratory tests in conjunction with ELISA techniques to evaluate atherogenic and lipid-related damage biomarkers. As displayed in ESF, Table 4, the dataset represented a wide geographical distribution, with United State, Turkey and Spain contributing the highest number of studies (4 studies for each country). Saudi Arabia, Poland, Russia and Germany each provided 3 studies, while Brazil, China and India contributed 3 each. Single contributions were noted from, Argentina, Australia, Austria, Chile, Egypt, Greece, Japan, Italy, Malta, Mexico, Oman, Romania, Slovakia, and Ukraine.

**Table 4.**
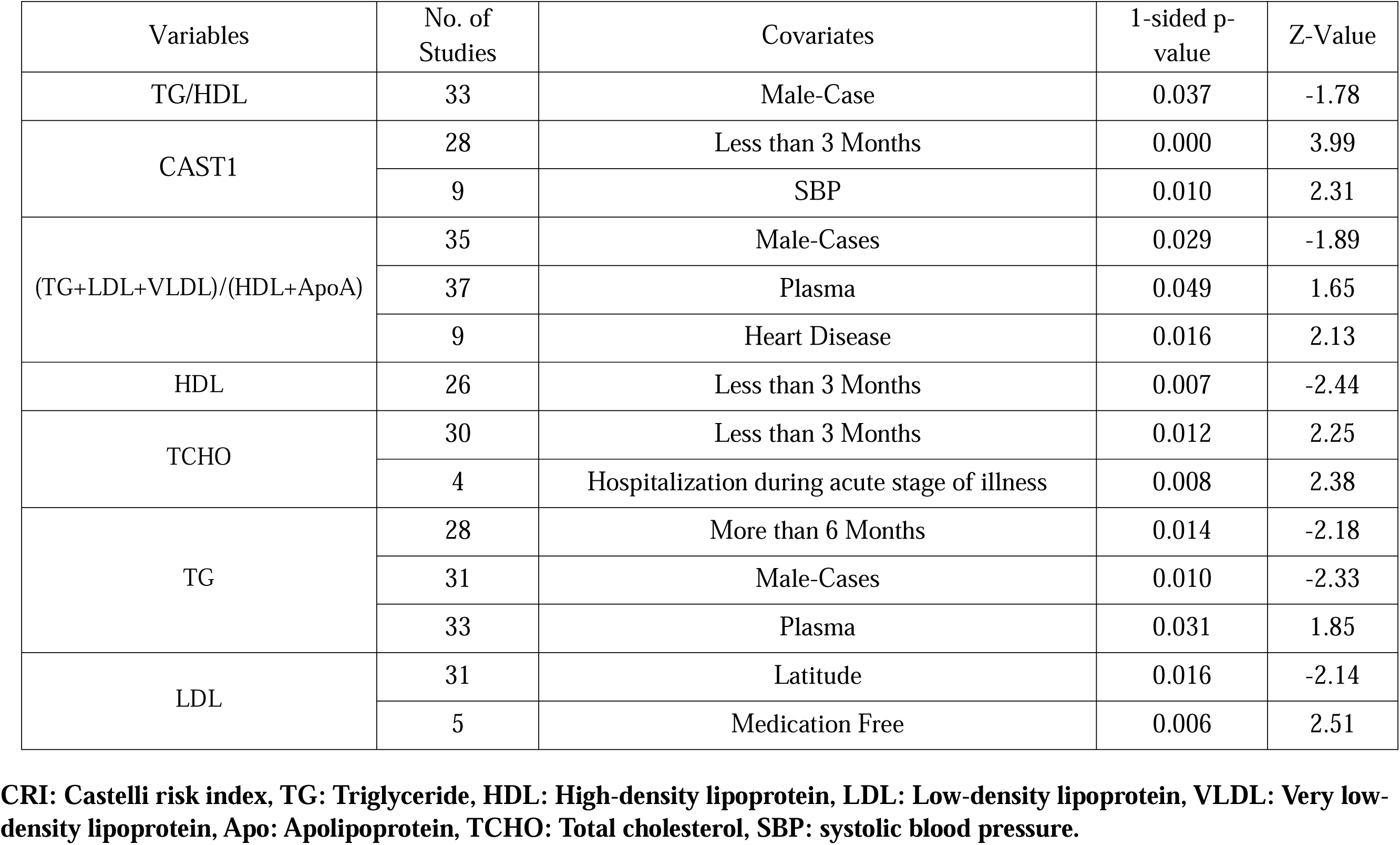
Results of Meta-regression.

Study quality and methodological robustness were evaluated, yielding median values of 4 (range: 2–7) for quality scores and 17 (range: 6–16) for redpoints. Detailed results are summarized in ESF Table 4.

### Primary Outcome Variables

#### CRI1 and CRI2 in Long COVID Patients vs. Normal Controls

The effect size for CRI1, derived from 38 studies, is summarized in **Table 1** and illustrated in **Figure 2**. One study reported CIs entirely below zero, while 10 studies reported CIs entirely above zero. The remaining 27 studies had overlapping CIs, with 11 showing negative SMD values and 16 reporting positive SMD values. A significant increase in CRI1 was observed in LC patients compared to NC (Table 2 and Figure 2). Publication bias analysis, summarized in **Table 3**, showed no significant evidence of bias. The effect size for CRI2, calculated from 33 studies, is detailed in Table 1 and **Figure 3**. Among these studies, one reported CIs entirely below zero, while nine reported CIs entirely above zero. The remaining 23 studies exhibited overlapping CIs, with eight showing negative SMD values and 15 reporting positive SMD values. Table 2 and Figure 3 indicate a significant elevation in CRI2 among LC patients compared to NC. The publication bias analysis, outlined in Table 3, revealed no significant bias.

**Figure 2.**
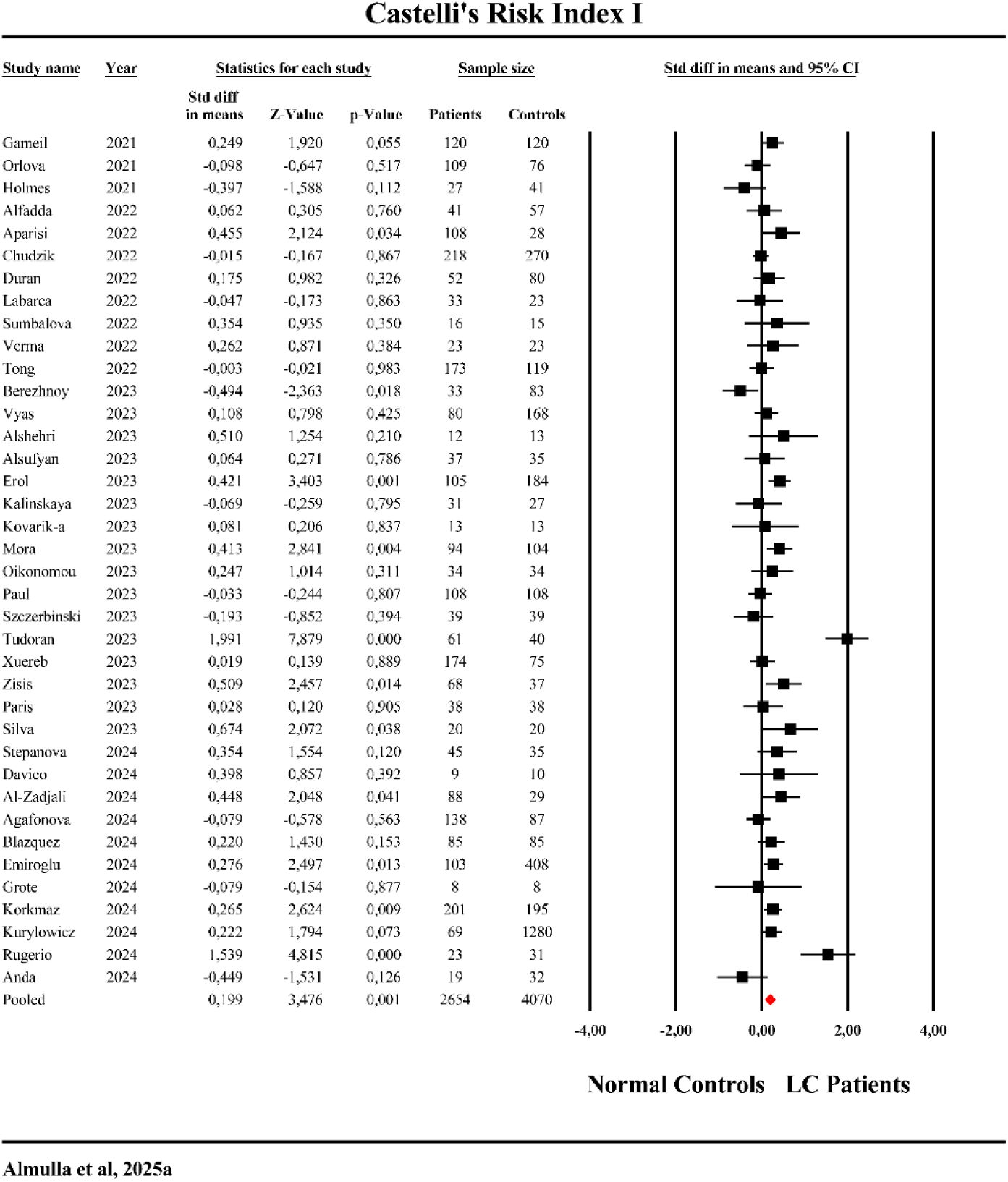
The forest plot of Castelli Risk Index-1 in patients with Long COVID (LC) and normal controls.

**Figure 3.**
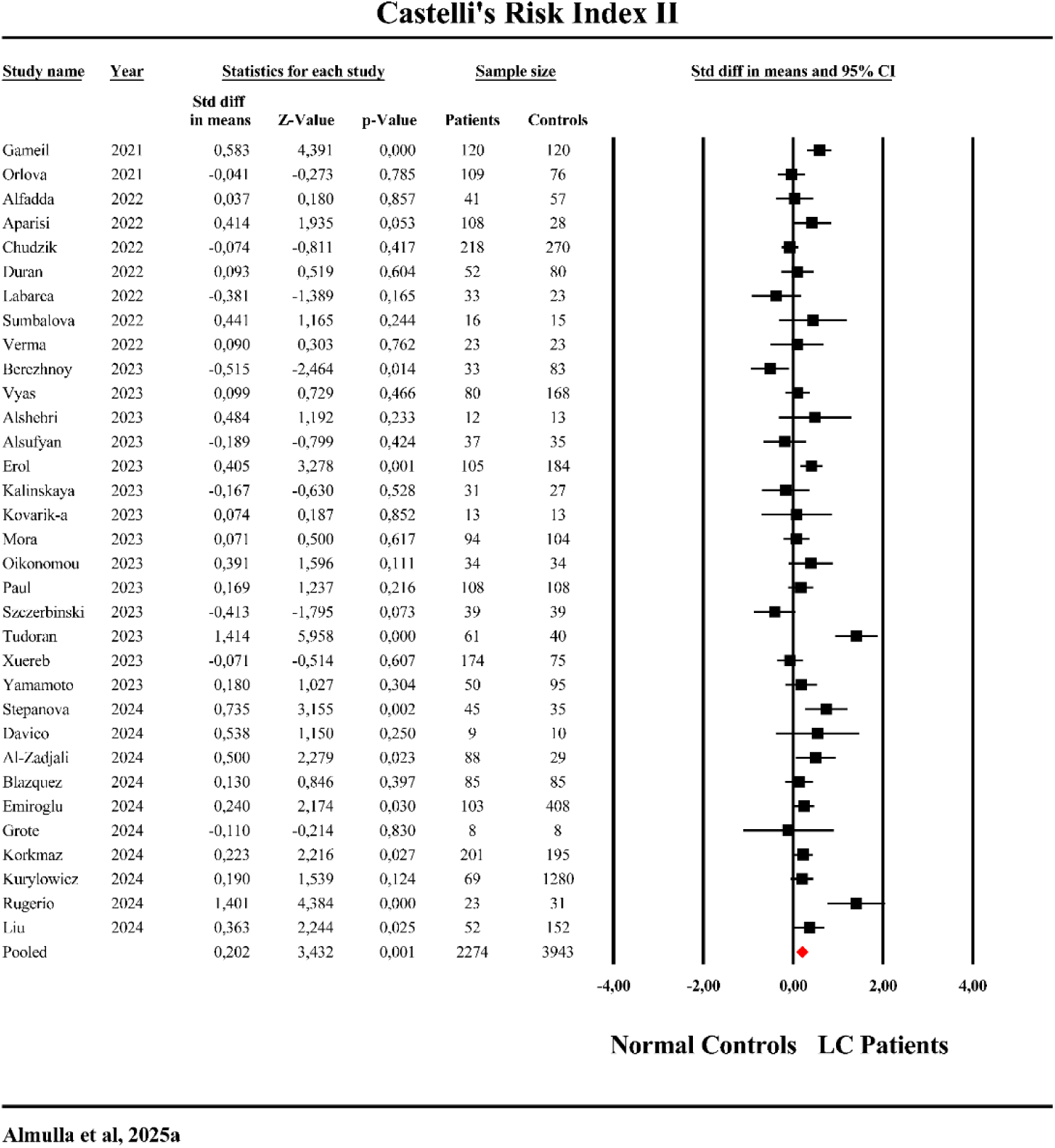
The forest plot of Castelli Risk Index-2 in patients with Long COVID (LC) and normal controls.

#### TG/HDL, (TG+LDL+VLDL)/(HDL+ApoA), and ApoB/ApoA Ratios

The TG/HDL ratio effect size, derived from 37 studies, is summarized in Table 1 and depicted in **Figure 4**. Two studies reported CIs entirely below zero, indicating a negative effect, while 13 studies reported CIs entirely above zero, indicating a positive effect. The remaining 22 studies had overlapping CIs, with seven reporting negative SMD values and 15 showing positive SMD values. Table 2 and Figure 2 show a significant increase in the TG/HDL ratio in LC patients compared to NC. No publication bias was detected as shown in Table 3.

**Figure 4.**
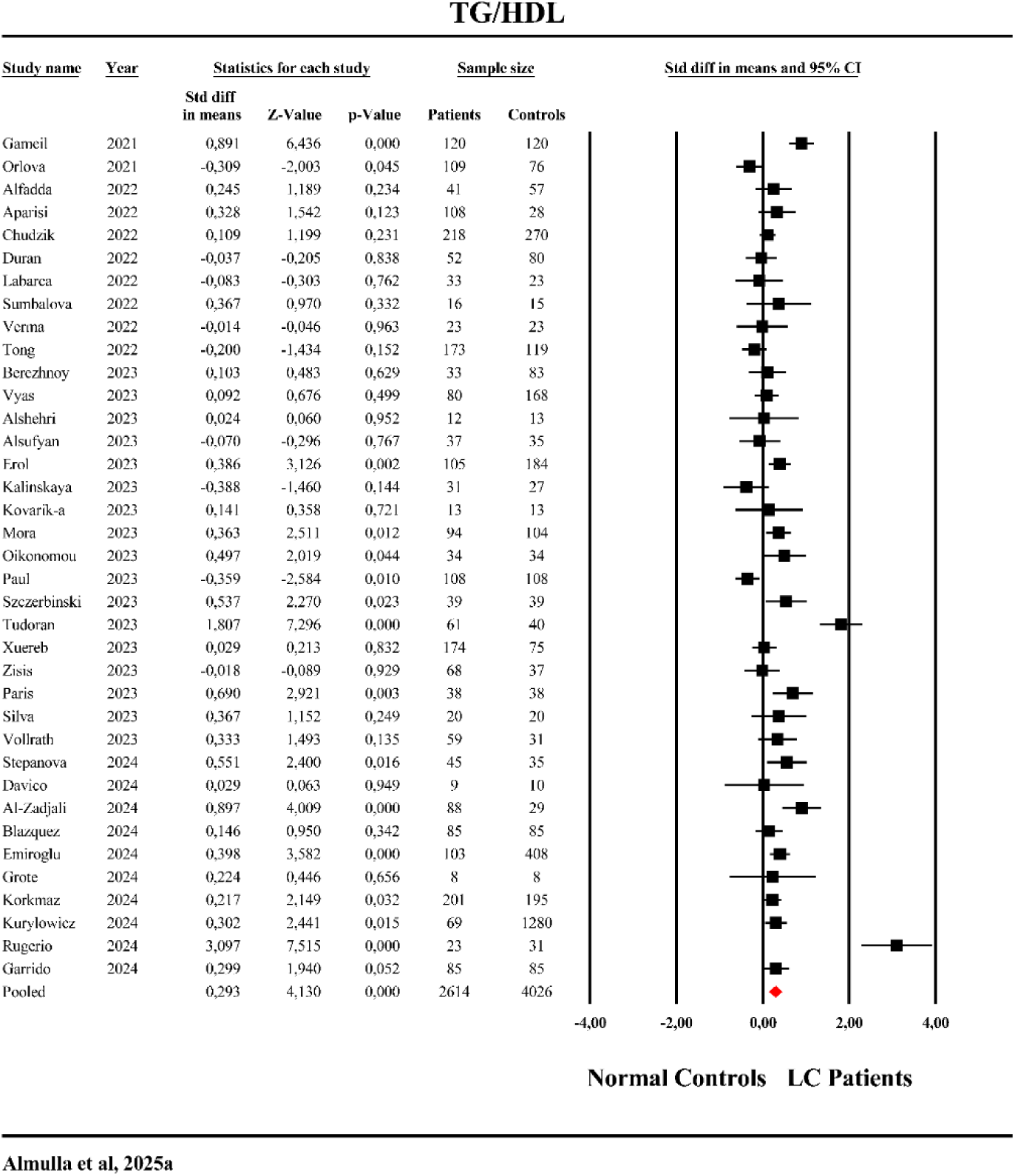
The forest plot of triglyceride (TG)/high-density lipoprotein (HDL) in patients with Long COVID (LC) and normal controls.

The effect size for the (TG+LDL+VLDL)/(HDL+ApoA) ratio, based on 39 studies, is summarized in Table 1 and illustrated in **Figure 5**. One study reported CIs entirely below zero, indicating a negative effect, while 12 studies presented CIs entirely above zero, reflecting a positive effect. The remaining 24 studies exhibited overlapping CIs, with nine showing negative SMD values and 15 reporting positive SMD values. Table 2 and Figure 2 indicate a significant increase in this ratio in LC patients compared to NC. Publication bias analysis, as outlined in Table 3, identified no significant evidence of bias.

**Figure 5.**
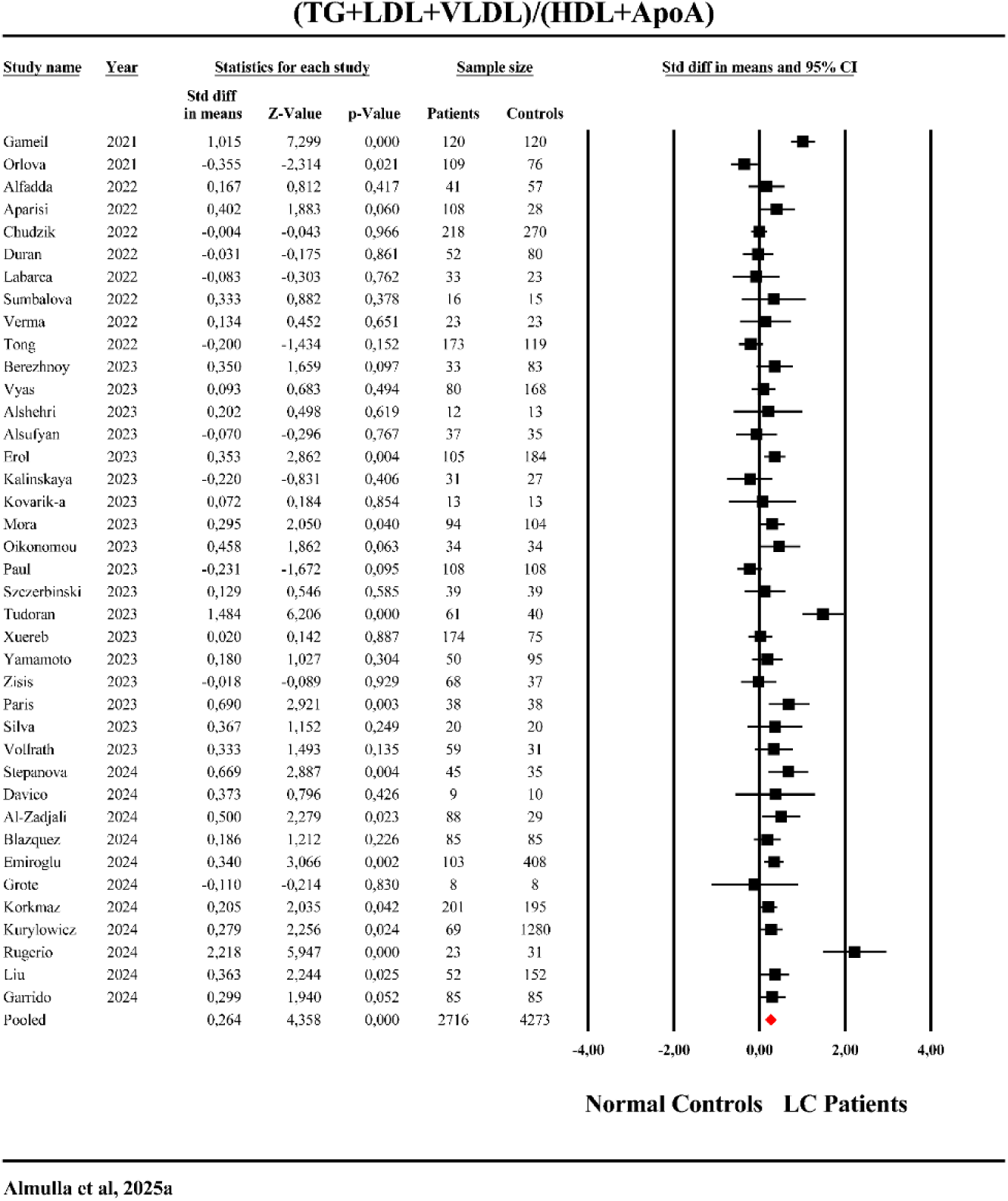
The forest plot of triglyceride (TG) + low-density lipoprotein (LDL)+very low-density lipoprotein (VLDL)/high-density lipoprotein (HDL) in patients with Long COVID (LC) and normal controls.

The effect size for the ApoB/ApoA ratio, derived from 3 studies, is summarized in Table 1 and ESF, Figure 1. None of the studies reported CIs entirely below zero; two showed CIs above zero, while one displayed overlapping CIs with a negative SMD value. Table 2 highlight a significant elevation in this ratio in LC patients compared to NC. Egger’s and Kendall tests indicated no significant evidence of bias.

### Secondary Outcome Variables

#### HDL

The effect size for HDL, derived from 31 studies, is presented in Table 1 and **Figure 6**. Eight studies reported CIs entirely below zero, indicating a negative effect, while one showed CIs entirely above zero, suggesting a positive effect. The remaining 22 studies had overlapping CIs, with 14 showing negative SMD values and eight reporting positive SMD values. Table 2 and Figure 6 demonstrate a significant decrease in HDL levels in LC patients compared to NC. Table 3 revealed no significant bias.

**Figure 6.**
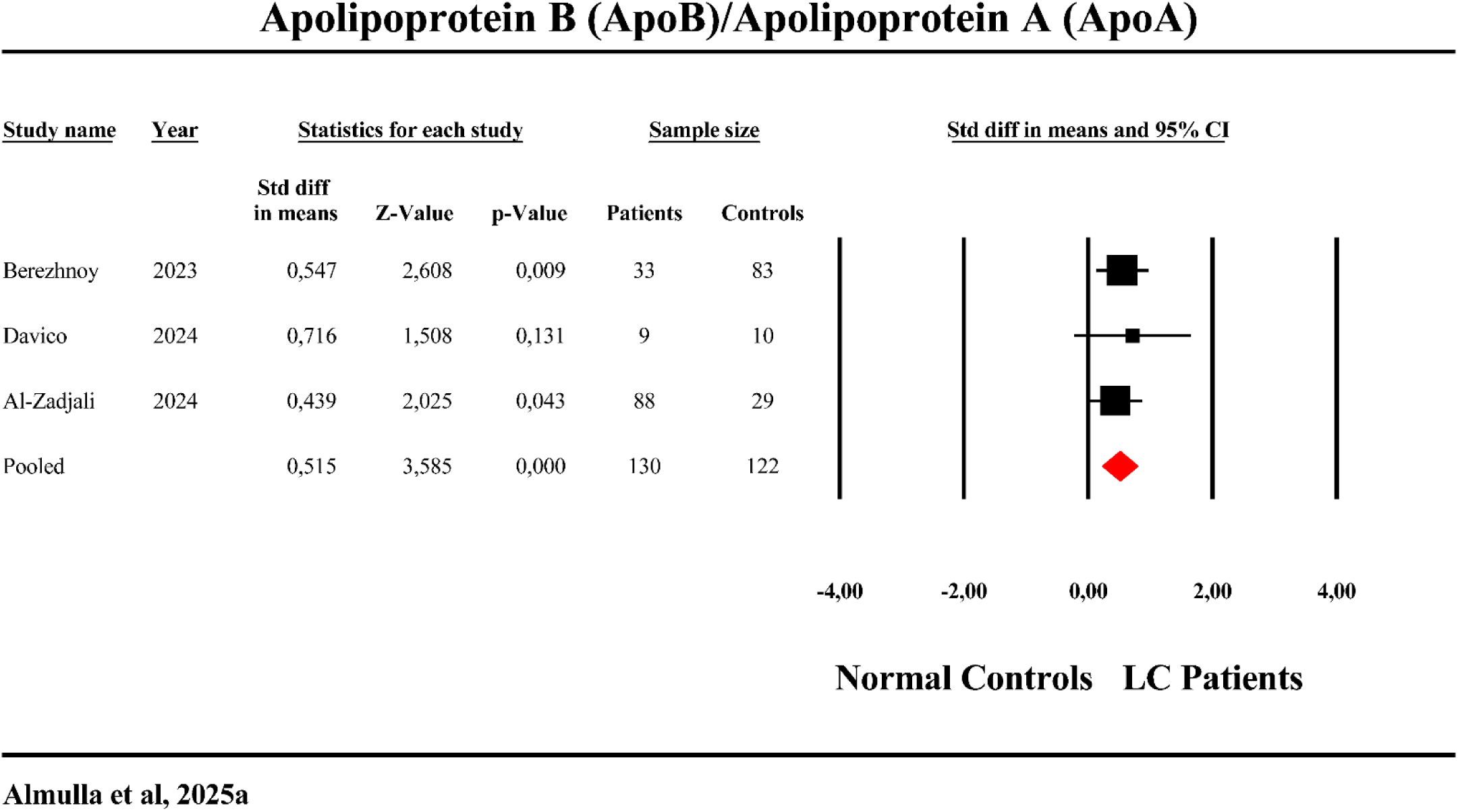
The forest plot of apolipoprotein (Apo)B/ApoA in patients with Long COVID (LC) and normal controls.

#### TG

The TG effect size, calculated from 31 studies, is detailed in Table 1 and Figure 2. Two studies reported CIs entirely below zero, suggesting a negative effect, while 13 studies showed CIs entirely above zero, indicating a positive effect. The remaining 20 studies displayed overlapping CIs, with six showing negative SMD values and 14 reporting positive SMD values. Table 2 and Figure 2 highlight a significant elevation in TG levels in LC patients compared to NC. No significant evidence of bias was observed (see Table 3).

#### TCHO

The effect size for TCHO, derived from 34 studies, is summarized in Table 1 and ESF, Figure 2. Among these, one study reported CIs entirely below zero, indicating a negative effect, while 10 studies showed CIs entirely above zero, reflecting a positive effect. The remaining 23 studies exhibited overlapping CIs, with 11 reporting negative SMD values and 12 showing positive SMD values. Table 2 and ESF, Figure 2 show a significant increase in TCHO levels in LC patients compared to NC. Publication bias analysis revealed no significant evidence of bias (see Table 3).

#### LDL

The effect size for LDL, based on 31 studies, is summarized in Table 1 and illustrated in ESF, Figure 3. Three studies reported CIs entirely below zero, suggesting a negative effect, while seven studies had CIs entirely above zero, reflecting a positive effect. The remaining 21 studies displayed overlapping CIs, with four showing negative SMD values and 17 reporting positive SMD values. Table 2 and ESF, Figure 3 demonstrate a significant elevation in LDL levels in LC patients compared to NC. Table 3 showed no significant bias.

#### VLDL

The effect size for VLDL, derived from 4 studies, is summarized in Table 1 and illustrated in ESF, Figure 4. Table 2 and ESF, Figure 4 indicates no significant difference in VLDL levels between LC patients and NC. Table 3 revealed no significant bias.

#### ApoB

The effect size for ApoB, based on three studies, is summarized in Table 1 and ESF, Figure 5. None of the studies reported CIs entirely below zero, while one showed CIs entirely above zero, indicating a positive effect. The other two studies displayed overlapping CIs, with both reporting positive SMD values. Table 2 and ESF, Figure 5 show a significant increase in ApoB levels in LC patients compared to NC. Table 3 indicates no significant evidence of bias.

#### ApoA

The ApoA effect size, derived from two studies, is summarized in Table 1 and illustrated in ESF, Figure 6. One study reported CIs entirely below zero, while none presented CIs entirely above zero. The remaining study exhibited overlapping CIs, with both studies reporting negative SMD values. Table 2 and ESF, Figure 6 reveal a significant reduction in ApoA levels in LC patients compared to NC. Table 3 showed no significant bias.

#### Meta-regression analysis

A meta-regression analysis (ESF, Table 5) was conducted to identify factors contributing to the heterogeneity observed in studies assessing atherogenic biomarkers in LC patients versus normal controls. The duration of the post-COVID period emerged as a significant factor, with durations under three months influencing CRI1, HDL, and TCHO levels, while durations exceeding six months impacted TG levels. Male gender was also identified as a significant contributor, affecting outcomes such as TG/HDL, (TG+LDL+VLDL)/(HDL+ApoA) ratios, and TG levels. Additional variables, including plasma medium, geographic latitude, absence of medication, hospitalization during the acute phase, elevated systolic blood pressure, and quality control measures, were found to significantly influence some outcomes, as outlined in ESF, Table 5.

## Discussion

### Atherogenicity in LC

This study is the first systematic review and meta-analysis to comprehensively evaluate atherogenicity indices and lipid profiles in LC patients compared to controls. The first major finding reveals that LC is associated with elevated CRI1, CRI2, atherogenic ratios, including TG/HDL and the comprehensive atherogenic index (TG+LDL+VLDL) / (HDL+ApoA). These shifts indicate an increase in the pro-atherogenic lipid profile and a decrease in anti-atherogenic capacity, suggesting an elevated risk for cardiovascular and metabolic complications including atherosclerosis and coronary artery disease. No previous studies have assessed CRI1, CRI2, or these atherogenic ratios in LC, though Stepanova et al. (2024) reported elevated atherogenic index of plasma (AIP) in LC patients.

Multiple interconnected mechanisms may elucidate lipid dysregulation and the elevated risk of cardiovascular disease in LC disease, including systemic inflammation, immune dysregulation, and oxidative stress (Papoutsidakis, Deftereos et al. 2014, Almulla, Thipakorn et al. 2023, Jorgensen, Macpherson et al. 2023). Elevated IL-6 and TNF-α in LC impair lipoprotein lipase (LPL) activity and enhance hepatic lipase activity, disrupting lipid metabolism (Chen, Xun et al. 2009, Kusunoki, Tsutsumi et al. 2013). Studies confirm increased lipase activity in LC patients (de Oliveira et al., 2022; Kovarik et al., 2023). TNF-α further drives lipid accumulation in hepatocytes via the α AMPK/mTOR/SREBP-1 pathway (Lv et al., 2015). Our recent findings suggest that persistent liver dysfunction in LC, marked by elevated liver damage indices and transaminase activities, interacts with immune-inflammatory pathways, amplifying lipid dysregulation and reducing anti-atherogenic potentials. This highlights the liver’s central role in lipid metabolism and cardiovascular health in LC (Almulla, Thipakorn et al. 2024c).

Oxidative and nitrosative stress exacerbate lipid abnormalities in LC, leading to the generation of lipid oxidative products, including oxidized low-density lipoprotein (ox-LDL) and other peroxidation byproducts such as malondialdehyde (Al-Hakeim, Al-Rubaye et al. 2023, Zisis, Durieux et al. 2023). Reactive oxygen and nitrogen species (RONS) impair HDL functionality, triggering autoimmune processes via neoepitope formation and exacerbating endothelial dysfunction (Almulla, Thipakorn et al. 2023). These processes may also involve ferroptosis, an iron-dependent form of cell death that contributes to neuropsychiatric and cardiovascular complications in LC disease (Sousa, Yehia et al. 2023, Yang, Wu et al. 2023).

Acute COVID-19 studies have similarly identified elevated CRI1, CRI2 and an atherogenic index of plasma (TG/HDL) linked to severe outcomes, including myocardial damage and increased mortality (Turgay Yıldırım and Kaya 2021, Gunay-Polatkan, Caliskan et al. 2024). Our findings extend these observations, revealing persistent lipid dysfunction in LC patients 3–6 months post-recovery. This prolonged dyslipidemia likely perpetuates systemic inflammation and metabolic dysregulation, complicating recovery (Akhmedov 2023, Ahmed and Ahmed 2024). The potential reversibility of these lipid abnormalities with targeted interventions remains unclear and should be a focus of future studies. Dyslipidemia in LC also contributes to endothelial damage and coagulopathy. Secretory phospholipase A2 metabolizes phospholipids into pro-coagulant and pro-inflammatory mediators, exacerbating thrombosis (Farooqui, Farooqui et al. 2023, Goracci, Petito et al. 2023). Microthrombi and vascular inflammation further elevate cardiovascular risks (Xiang, Wu et al. 2023). The observed dyslipidemia may also enhance thrombotic events through altered lipoprotein composition, underscoring the need for early lipid and inflammatory marker monitoring.

The interplay between lipid metabolism and immune responses highlights the dual role of metabolic and inflammatory pathways in atherosclerosis (Gisterå and Ketelhuth 2018, Pirillo, Bonacina et al. 2018). In LC, activated immune-inflammatory responses, characterized by abnormal T-cell activity, likely contribute to the observed atherogenic lipid profiles (Almulla, Thipakorn et al. 2024a). These findings highlight the necessity of long-term monitoring and interventions to address the atherogenicity and cardiovascular implications of LC. Strategies that integrate the management of lipid dysregulation and inflammation are crucial for mitigating cardiovascular risks in LC.

### Hypercholesterolemia and hypertriglyceridemia in Long COVID

The second major finding of this study indicates significantly elevated levels of TCHO and TGs in LC patients compared to controls. Consistent with prior studies (Emiroglu, Dicle et al. 2024, Oliván-Blázquez, Bona-Otal et al. 2024), this dyslipidemic profile underscores the elevated atherogenicity in LC. Interestingly, acute COVID is characterized by decreased total TCHO and unchanged TG levels (Mahat, Rathore et al. 2021, Zinellu, Paliogiannis et al. 2021, Chidambaram, Shanmugavel Geetha et al. 2022).

Persistent inflammation and liver dysfunction likely underpin this metabolic shift in acute and LC patients (Almulla, Thipakorn et al. 2024a, Almulla, Thipakorn et al. 2024c). However, during acute COVID, severe inflammation and vascular leakage result in cholesterol efflux into alveolar spaces which probably reducing plasma levels (Wei, Zeng et al. 2020). In contrast, LC-associated chronic inflammation hinders hepatic cholesterol clearance and facilitates TG release, resulting in their accumulation in plasma (Al-Hakeim, Al-Rubaye et al. 2023, Maes, Almulla et al. 2023).

Elevated cholesterol levels augment immune responses by activating TLRs, inflammasomes, and pro-inflammatory pathways, increasing leukocyte production and systemic inflammation (Tall and Yvan-Charvet 2015). Al-Hakeim et al reported that LC-associated activation of the NLRP3 inflammasome, characterized by elevated IL-1β 18, and caspase-1 levels, contributes to neurotoxicity and systemic immune dysregulation (Al-Hakeim, Al-Rubaye et al. 2023). These findings highlight the bidirectional relationship between lipid metabolism and immune activation in LC.

LC is characterized by significant elevations in oxidative and nitrosative stress (Al-Hakeim, Al-Rubaye et al. 2023), which may promote the oxidation of cholesterol into oxysterols, compounds with notable neurotoxic potential. Oxysterols can cross the blood-brain barrier (BBB), where they activate microglia and exacerbate neuroinflammation (BjÖRkhem 2006, Gamba, Testa et al. 2015). Elevated cholesterol also weakens BBB integrity, facilitating immune cell infiltration into the central nervous system (CNS) (Rapp et al., 2008; de Oliveira et al., 2020). These mechanisms provide a plausible link between dyslipidemia and neurocognitive impairments frequently observed in LC (Almulla, Al-Hakeim et al. 2023).

Moreover, hypertriglyceridemia in LC adds complexity by enhancing immune-inflammatory responses via saturated fatty acid (SFA) metabolism to ceramide, activating mitogen-activated protein kinases (MAPKs), and increasing cytokine production (Schwartz, Zhang et al. 2010). Elevated TG levels also stimulate B-cell secretion of immunoglobulin M (IgM), a mechanism linked to autoimmunity, particularly in males (Shi, Guo et al. 2015). TGs accumulate in macrophages upon inflammatory activation which is mediated by hypoxia-inducible lipid droplet–associated (HILPDA) protein (van Dierendonck, Vrieling et al. 2022). Chronic immune activation in LC, characterized by α, may further drive TGs accumulation through hepatic synthesis and impaired clearance (Feingold and Grunfeld 1992). Additionally, mitochondrial dysfunction in LC impairs fatty acid β accumulation (Vankoningsloo, Piens et al. 2005, Molnar, Lehoczki et al. 2024) (Vankoningsloo et al., 2005; Molnar et al., 2024).

Furthermore, latent viral reactivation, particularly Epstein-Barr virus (EBV) and human herpesvirus 6 (HHV-6) in LC (Maes, Almulla et al. 2024, Vojdani, Almulla et al. 2024), may exacerbate lipid dysregulation. In this context, EBV upregulates LDL receptor expression, enhancing cholesterol uptake and viral replication (Wang, Wang et al. 2019), while HHV-6 depends on cholesterol for infectivity (Huang, Li et al. 2006). Peroxisome proliferator-activated receptor alpha, a regulator of cholesterol metabolism, also influences herpesvirus latency and reactivation (Reese, Tao et al. 2018), linking viral pathogenesis to dyslipidemia and inflammation.

### Lipoproteins and apolipoproteins profiles in LC

The study’s third major finding is that LC patients show significantly lower levels of HDL and ApoA, while exhibiting higher levels of LDL, ApoB, and the ApoB/ApoA ratio, in comparison to the NC group. These findings align with prior studies highlighting similar abnormalities in lipoprotein and apolipoprotein profiles in LC (Alshehri, AlQahtani et al. 2023, Berezhnoy, Bissinger et al. 2023, Erol, Tezcan et al. 2023, Davico, Martín et al. 2024). Notably, during the acute stage of COVID infection, lowered levels of HDL, LDL and ApoA were observed (Li, Zhang et al. 2021).

As mentioned earlier, reduced HDL and ApoA levels primarily influence RCT, limiting cholesterol clearance from peripheral tissues to the liver. This increase in lipid peroxidation leads to the accumulation of oxidized lipids, such as oxidized LDL and HDL (oxLDL and oxHDL), 4-hydroxynonenal, and malondialdehyde, which exacerbate oxidative stress and inflammation (Almulla, Thipakorn et al. 2023). These probably promote autoimmune reactions targeting neoepitopes and self-proteins as detected in LC (Almulla, Thipakorn et al. 2023, Almulla, Maes et al. 2024d). Notably, SARS-CoV-2 spike protein interactions with HDL particles may exacerbate lowered RCT and elevated atherogenicity by impairing HDL functionality, further amplifying lipid imbalances (Barter, Nicholls et al. 2004, Correa, Del Giudice et al. 2023). Dysfunctional HDL reduces its anti-inflammatory and atheroprotective capacities, allowing increased immune activation marked by elevated CRP, IL-6, and IL-17 (Stepanova, Driianska et al. 2024). Additionally, failure of HDL to inhibit platelet activation and the expression of endothelial adhesion molecules aggravates inflammation and thrombosis (Murphy and Woollard 2010, Haas and Mooradian 2011).

Increased LDL levels in LC present considerable cardiovascular risks. LDL promotes endothelial dysfunction and activates inflammatory pathways such as NF-Bκ and PI3K/AKT/mTOR, leading to oxidative stress (leading to formation highly immunogenic form of LDL namely oxLDL) and foam cell formation (Cominacini, Pasini et al. 2000, Rudijanto 2007, Zhang, Han et al. 2024). These processes are also linked to persistent immune activation in LC, as oxLDL interacts with monocytes and macrophages through scavenger receptors like LOX-1 and TLRs, inducing pro-inflammatory cytokines and adhesion molecule expression (Choi, Yin et al. 2013, Xu, Xiwen et al. 2019). This underscores a direct connection between lipid peroxidation, chronic inflammation, and vascular damage in LC.

ApoA’s anti-inflammatory and antioxidant properties are critical for modulating immune cell activation and cytokine production via pathways like JAK2/STAT3 (Navab et al., 2005; Yin et al., 2011). Reduced ApoA levels in LC patients may compromise these protective effects, contributing to sustained systemic inflammation and oxidative stress seen in LC. Additionally, anti-Apo A-1 antibodies (AAA1), linked to persistent respiratory symptoms in COVID-19 patients, suggest a potential role in LC symptomatology (L’Huillier, Pagano et al. 2022).

Conversely, elevated ApoB levels and ApoB/ApoA ratios are markers of atherogenicity and correlate strongly with cardiovascular disease severity (Tabas, Williams et al. 2007, Kraaijenhof, Hovingh et al. 2021). Modifications of ApoB in LDL particles enhance their retention in arterial walls, triggering maladaptive immune responses that propagate lesion formation (Malekmohammad, Bezsonov et al. 2021). Future research should explore therapeutic strategies aimed at restoring HDL function, lowering LDL and ApoB levels, and mitigating inflammation to reduce the long-term cardiovascular and metabolic risks associated with LC.

Statins, widely used cholesterol-lowering drugs known to elevate HDL levels, exhibit anti-inflammatory properties that alleviate symptoms of LC. Their combination with angiotensin II type 1 receptor blockers (ARBs) has been shown to enhance clinical outcomes and reduce the inflammatory potential of the HDL proteome in LC patients (Grote, Schaefer et al. 2024). Other treatments proposed for LC include therapeutic apheresis, since this modifier of the lipid profile can decrease inflammatory mediators and improve the clinical symptoms (Jaeger, Arron et al. 2022, Achleitner, Steenblock et al. 2023). Targeted interventions are needed regarding inflammation-anti-IL-6 therapy, oxidative stress (such as N-acetylcysteine), and triglycerides (for example, fibrates and omega-3 fatty acids)-along with lifestyle changes to mitigate risks and improve clinical outcomes among LC patients (Yang, Chang et al. 2022, Bradbury, Wilkinson et al. 2023, Barlattani, Celenza et al. 2024, Simonetti, Restaino et al. 2024).

## Limitations

The study demonstrated no evidence of publication bias across the measured outcomes, as confirmed by non-significant Egger’s and Kendall’s tests. However, several limitations should be considered. The analysis was restricted to specific aspects of the RCT, as data on paraoxonase 1 (PON1) and lecithin-cholesterol acyltransferase (LCAT), key components associated with immune responses, were unavailable due to a lack of studies examining their levels in LC patients versus controls. This gap limited the study’s ability to comprehensively assess RCT mechanisms in LC. Additionally, while meta-regression analysis revealed no significant impact of body mass index (BMI) on the outcomes, insufficient reporting of metabolic syndrome in the included studies prevented an evaluation of its influence on lipid profiles in LC patients. This is particularly important as metabolic syndrome has been shown to confound lipid abnormalities in related conditions, such as major depression (Maes, Jirakran et al. 2024, Maes, Vasupanrajit et al. 2024). Moreover, the study could not investigate the effects of medications administered to LC patients due to the limited number of studies reporting such information, which may have influenced lipid and immune profiles. Addressing these limitations in future research could enhance understanding of the complex relationships between lipid metabolism, immune responses, and clinical factors in LC.

## Conclusions

The findings of the current study are summarized in **Figure 7**, which indicates that LC is linked to increased atherogenicity and reduced anti-atherogenic potential, confirming its role as a pro-atherogenic condition that may lead to serious cardiovascular complications. The findings underscore the significance of routine lipid profiling in individuals with LC to detect and manage dyslipidaemia early. Timely implementation of lipid-lowering therapies is crucial for mitigating atherogenic progression and decreasing the risk of cardiovascular events. These results necessitate further investigation into the mechanisms of lipid dysregulation in LC and the development of targeted therapeutic strategies to enhance patient outcomes and long-term cardiovascular health.

**Figure 7.**
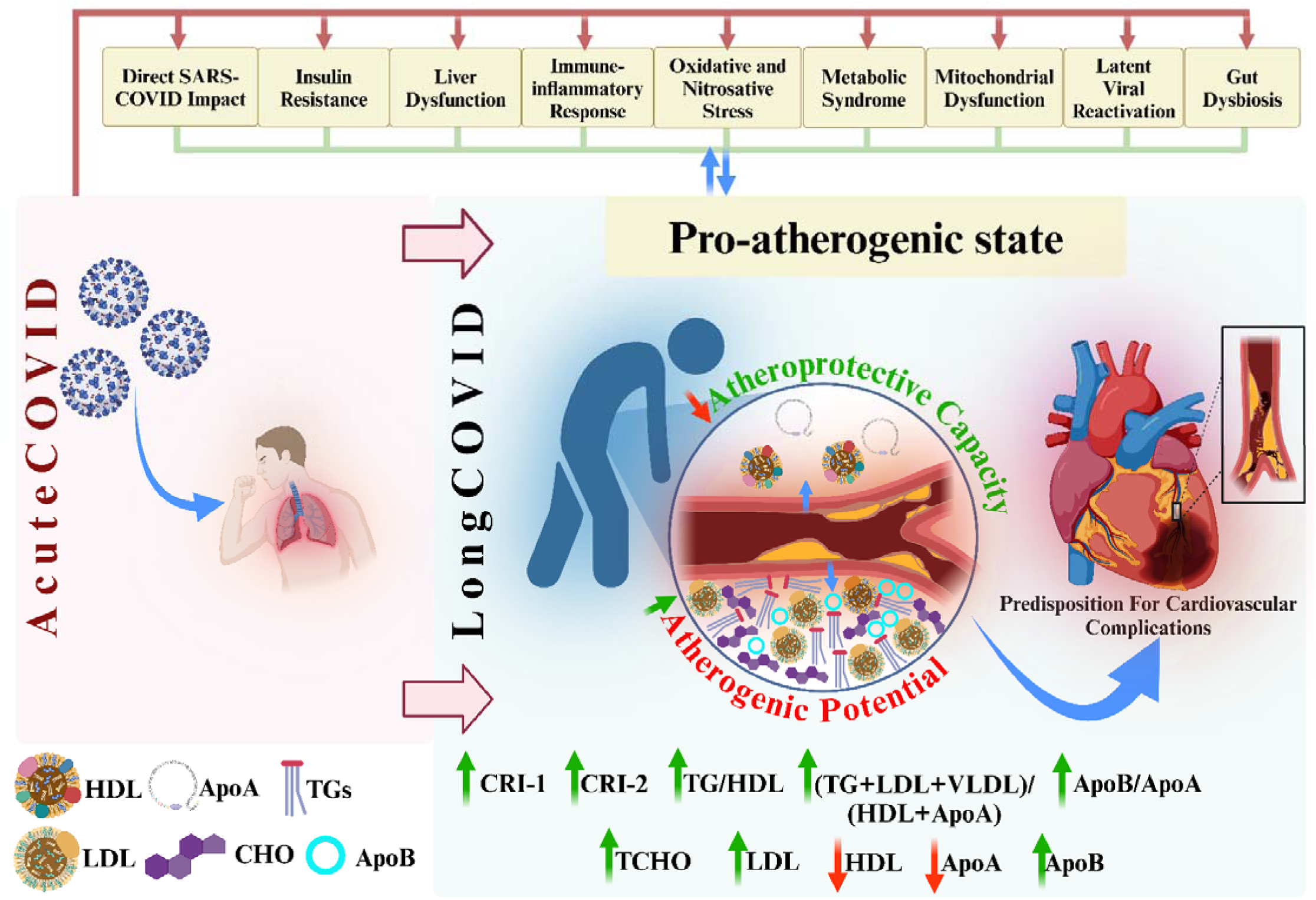
Summary of the elevated atherogenicity in Long COVID (LC) disease. HDL: High-density lipoprotein, Apo: Apolipoprotein, TGs: Triglycerides, LDL: Low-density lipoprotein, CHO: Cholesterol, CRI: Castelli risk index, VLDL: very low-density lipoprotein.

## Ethical approval and consent to participate

Not applicable.

## Consent for publication

Not applicable.

## Availability of data and materials

The corresponding author (MM) will consider reasonable requests for access to the dataset (Excel file) utilized in this meta-analysis, following the completion of data usage by all contributing authors.

## Author’s contributions

AA and MM conceptualized and designed the study. Data collection was conducted collaboratively by AA and YT. Statistical analyses were performed by AA and MM. All authors contributed to drafting the manuscript, reviewed it critically for important intellectual content, and approved the final version for submission.

## Acknowledgments

Not applicable.

