## supplementary file for "Elevated Atherogenicity in Long COVID: A Systematic Review and Meta-Analysis"

**SHORT TITLE:** Atherogenic Indices and Lipid Dysregulation in Long COVID

(1-3) Abbas F. Almulla, (4) Yanin Thipakorn, (1,2,4,5-8) Michael Maes

1. Sichuan Provincial Center for Mental Health, Sichuan Provincial People's Hospital, School of Medicine, University of Electronic Science and Technology of China, Chengdu 610072, China

2. Key Laboratory of Psychosomatic Medicine, Chinese Academy of Medical Sciences, Chengdu 610072, China

3. Medical Laboratory Technology Department, College of Medical Technology, The Islamic University, Najaf, Iraq

4. Department of Psychiatry, Faculty of Medicine, Chulalongkorn University and King Chulalongkorn Memorial Hospital, the Thai Red Cross Society, Bangkok, Thailand.

5. Cognitive Fitness and Technology Research Unit, Faculty of Medicine, Chulalongkorn University, Bangkok, Thailand.

6. Department of Psychiatry, Medical University of Plovdiv, Plovdiv, Bulgaria.

7. Research Institute, Medical University Plovdiv, Plovdiv, Bulgaria.

8. Kyung Hee University, 26 Kyungheedae-ro, Dongdaemun-gu, Seoul 02447, Republic of Korea.

**Corresponding author:**

Prof. Dr. Michael Maes, M.D., Ph.D.

Sichuan Provincial Center for Mental Health

Sichuan Provincial People’s Hospital,

School of Medicine,

University of Electronic Science and Technology of China

Chengdu 610072

China

[Michael Maes Google Scholar profile](https://scholar.google.com/citations?user=1wzMZ7UAAAAJ&hl=en)

<https://scholar.google.co.th/citations?user=1wzMZ7UAAAAJ&hl=th&oi=ao>

Highly cited author: 2003-2023 (ISI, Clarivate)

ScholarGPS: Worldwide #1 in molecular neuroscience; #1/4 in pathophysiology

Expert worldwide medical expertise ranking, Expertscape (December 2022), worldwide:

#1 in CFS, #1 in oxidative stress, #1 in encephalomyelitis, #1 in nitrosative stress, #1 in nitrosation, #1 in tryptophan, #1 in aromatic amino acids, #1 in stress (physiological), #1 in neuroimmune; #2 in bacterial translocation; #3 in inflammation, #4-5: in depression, fatigue and psychiatry.

**ESF. Table 1.** Search sentences and terms were used in each database.

| **Database Name** | **Search Sentence** | **No. of Articles** |
| --- | --- | --- |
| **PubMed/Medline** | ("Long COVID" OR "Post-COVID syndrome" OR "Post-Acute COVID-19 Syndrome"[MeSH] OR "PASC") AND ("Lipid profile" OR "Lipids"[MeSH] OR "Dyslipidemia" OR "Cholesterol"[MeSH] OR "Triglycerides"[MeSH] OR "HDL-C" OR "LDL-C" OR "Apolipoproteins"[MeSH]) AND ("Atherogenic index" OR "Atherogenic biomarkers" OR "Atherogenic lipid profile" OR "Oxidized LDL" OR "Lipoprotein(a)" OR "Lipoproteins"[MeSH]) | **20** |
|  | (("Long COVID" OR "Post-COVID syndrome" OR "Post-acute sequelae of SARS-CoV-2" OR "Chronic COVID" OR "PASC") AND ("Lipid profile" OR "Lipids"[MeSH] OR "Cholesterol"[MeSH] OR "Dyslipidemia" OR "HDL-C" OR "LDL-C" OR "Triglycerides"[MeSH]) AND ("Atherogenic index" OR "Atherosclerosis"[MeSH] OR "Oxidized LDL" OR "Lipoprotein(a)" OR "Apolipoproteins"[MeSH])) | **9** |
|  | (("lipid metabolism"[MeSH Terms] OR "dyslipidemia"[All Fields] OR "LDL"[All Fields] OR "HDL"[All Fields] OR "VLDL"[All Fields] OR "cholesterol"[All Fields] OR "triglycerides"[All Fields]) AND ("Long COVID"[All Fields] OR "PASC"[All Fields] OR "post-COVID syndrome"[All Fields]) AND ("alterations"[All Fields] OR "biomarkers"[All Fields] OR "changes"[All Fields]) AND ("adults"[All Fields] OR "COVID-19 survivors"[All Fields] OR "patients"[All Fields])) | **16** |
|  | (("Lipid profiles"[MeSH] OR "LDL"[All Fields] OR "HDL"[All Fields] OR "triglycerides"[All Fields] OR "cholesterol"[All Fields] OR "VLDL"[All Fields]) AND ("Long COVID"[All Fields] OR "post-acute sequelae of SARS-CoV-2 infection"[All Fields] OR "PASC"[All Fields]) AND ("alterations"[All Fields] OR "levels"[All Fields] OR "changes"[All Fields] OR "dyslipidemia"[All Fields] OR "biomarkers"[All Fields])AND ("cross-sectional studies"[All Fields] OR "case-control studies"[All Fields] OR "cohort studies"[All Fields])) | **2** |
| **Google Scholar** | ("cholesterol" OR "total cholesterol") AND ("Long COVID" OR "post-COVID syndrome" OR "post-acute sequelae of SARS-CoV-2 infection" OR "PASC") AND ("altered lipid profile" OR "lipid biomarkers" OR "dyslipidemia" OR "serum cholesterol levels") AND ("case-control study" OR "cohort study" OR "cross-sectional study" OR "primary research") AND ("COVID-19 patients" OR "adults") -review -meta-analysis | **946** |
|  | ("Triglycerides" OR "TG" OR "serum triglycerides") AND ("Long COVID" OR "post-COVID syndrome" OR "PASC") AND ("lipid abnormalities" OR "alterations in lipid levels" OR "dyslipidemia") AND ("case-control study" OR "clinical study" OR "cohort study") AND ("COVID-19 patients" OR "COVID-19 survivors") -review -meta-analysis | **4** |
|  | ("HDL" OR "high-density lipoprotein") AND ("Long COVID" OR "post-COVID syndrome" OR "post-acute sequelae of SARS-CoV-2 infection" OR "PASC") AND ("lipid metabolism" OR "biomarkers" OR "dyslipidemia" OR "alterations in lipid profile") AND ("clinical study" OR "primary research" OR "observational study") AND ("adults" OR "COVID-19 survivors") -review -meta-analysis | **452** |
|  | ("LDL" OR "low-density lipoprotein") AND ("Long COVID" OR "post-COVID syndrome" OR "post-acute sequelae of SARS-CoV-2 infection" OR "PASC") AND ("lipid profile" OR "dyslipidemia" OR "altered lipid metabolism" OR "changes in lipid levels") AND ("case-control study" OR "cohort study" OR "cross-sectional study" OR "observational study") AND ("adults" OR "COVID-19 survivors") -review -meta-analysis | **861** |
| **SciFinder** | ("LDL" OR "low-density lipoprotein" OR "HDL" OR "high-density lipoprotein" OR "triglycerides" OR "TG" OR "cholesterol" OR "VLDL") AND ("Long COVID" OR "post-acute sequelae of SARS-CoV-2 infection" OR "PASC" OR "post-COVID syndrome") AND ("alterations" OR "levels" OR "changes" OR "dyslipidemia" OR "biomarkers") AND ("cross-sectional studies" OR "case-control studies" OR "cohort studies") AND ("adults" OR "patients" OR "COVID-19 survivors") | **11** |
| **SCOPUS** | (TITLE-ABS-KEY ("Long COVID" OR "post-COVID syndrome" OR "post-acute sequelae of SARS-CoV-2 infection" OR "PASC") AND TITLE-ABS-KEY("lipid profile" OR "lipid markers" OR "lipid metabolism" OR "dyslipidemia" OR "LDL" OR "HDL" OR "triglycerides" OR "cholesterol" OR "VLDL")  AND TITLE-ABS-KEY("alterations" OR "levels" OR "changes" OR "biomarkers")  AND TITLE-ABS-KEY("patients" OR "COVID-19 survivors" OR "adults")) | **112** |

**ESF. Table 2.** Immune cofounder’s scale (ICS) applied from Andrés-Rodríguez. et al.. 2019

| **Methodological quality of the study** | |
| --- | --- |
| **1** | Study sample ≥ 128 participants including patients and controls (1= Yes. 0 = No) |
| **2** | Did the study control the results for potential confounders (e.g.. age. BMI. gender. race)? (1= Yes. 0 = No) |
| **3** | Were participants with Long COVID and controls age- and-gender-matched or was there a statistical control? (1= Yes. 0 = No) |
| **4** | Was the time of sample collection specified (e.g.. morning vs. evening)? (1= Yes. 0 = No) |
| **5** | Were participants with Long COVID free of immunomodulatory drugs including anti-cytokines. glucocorticoids. immunoglobulins. and immunosuppressants. or was there a medication washout period. or was drug intake statistically controlled for? (1= Yes. 0 = No) |
| **6** | Were participants with Long COVID free of antidepressants and mood stabilizers or were the data statistically controlled for? (1= Yes. 0 = No) |
| **7** | Reporting either the manufacturer of the test or detection limit and coefficients of variation (1= Yes. 0 = No) |
| **8** | Reporting how data under detection limit were handled (1 = Yes. 0 = No) |
| **9** | Reporting % of the sample under detection limit (1=Yes. 0= No) |
| **10** | Reporting blood fraction (serum. plasma. culture supernatant or whole blood) (1= Yes. 0 = No) |
| **Total quality score (10 points)** | |
| **Biomarker confounders red points**  *The red points should not be given if the item is statistically controlled for* | |
| **1** | 3 red points for comorbid illnesses such as autoimmune disorders & other immune disorders including rheumatoid arthritis. psoriasis. inflammatory bowel disease. chronic obstructive pulmonary disease. multiple sclerosis |
| **2** | 3 red points for use of recreational drugs such as methamphetamine or opioids |
| **3** | 2 red points when groups were not matched for age |
| **4** | 2 red points when groups were not matched for sex |
| **5** | 2 red points for medication use as for example immunomodulators |
| **6** | 2 red points for early traumatic life events |
| **7** | 2 red points for shift work and primary sleep disorders |
| **8** | 1.5 red points for use of antipsychotics |
| **9** | 1 red point for more common systemic immune disorders including diabetes type 1/2. essential hypertension. metabolic syndrome |
| **10** | 1 red point for not fasting (8 hours before blood extraction) |
| **11** | 1 red point for use of omega-3 and antioxidant supplements |
| **12** | 1 red point when data were not controlled for body mass index |
| **13** | 1 red point when data were not controlled for physical activity or sedentary life |
| **14** | 1 red point when data were not controlled for smoking |
| **15** | 1 red point for use of oral contraceptives or NSAIDs |
| **16** | 0.5 red points when data were not controlled for ethnicity in countries such as US. Brazil |
| **17** | 0.5 red points when data were not controlled for seasonality |
| **18** | 0.5 red points when data were not controlled for diurnal variation (8-10 a.m. versus all other time points) |
|  | **Total red point score (26 points)** |

**ESF. Table 3.** PRISMA checklist

| **Section/topic** | **#** | **Checklist item** | **Reported on page #** |
| --- | --- | --- | --- |
| **TITLE** | | | |
| Title | 1 | Identify the report as a systematic review. meta-analysis. or both. | 1 |
| **ABSTRACT** | | | |
| Structured summary | 2 | Provide a structured summary including. as applicable: background; objectives; data sources; study eligibility criteria. participants. and interventions; study appraisal and synthesis methods; results; limitations; conclusions and implications of key findings; systematic review registration number. | 3 |
| **INTRODUCTION** | | | |
| Rationale | 3 | Describe the rationale for the review in the context of what is already known. | 5 |
| Objectives | 4 | Provide an explicit statement of questions being addressed with reference to participants. interventions. comparisons. outcomes. and study design (PICOS). | 8 |
| **METHODS** | | | |
| Protocol and registration | 5 | Indicate if a review protocol exists. if and where it can be accessed (e.g.. Web address). and. if available. provide registration information including registration number. | 9 |
| Eligibility criteria | 6 | Specify study characteristics (e.g.. PICOS. length of follow-up) and report characteristics (e.g.. years considered. language. publication status) used as criteria for eligibility. giving rationale. | 10 |
| Information sources | 7 | Describe all information sources (e.g.. databases with dates of coverage. contact with study authors to identify additional studies) in the search and date last searched. | 9 |
| Search | 8 | Present full electronic search strategy for at least one database. including any limits used. such that it could be repeated. | ESF. Table 1. page 3 |
| Study selection | 9 | State the process for selecting studies (i.e.. screening. eligibility. included in systematic review. and. if applicable. included in the meta-analysis). | 10 |
| Data collection process | 10 | Describe method of data extraction from reports (e.g.. piloted forms. independently. in duplicate) and any processes for obtaining and confirming data from investigators. | 11 |
| Data items | 11 | List and define all variables for which data were sought (e.g.. PICOS. funding sources) and any assumptions and simplifications made. | 11 |
| Risk of bias in individual studies | 12 | Describe methods used for assessing risk of bias of individual studies (including specification of whether this was done at the study or outcome level). and how this information is to be used in any data synthesis. | 12 |
| Summary measures | 13 | State the principal summary measures (e.g.. risk ratio. difference in means). | 12 |
| Synthesis of results | 14 | Describe the methods of handling data and combining results of studies. if done. including measures of consistency (e.g.. I^2^) for each meta-analysis. | 12 |
| Risk of bias across studies | 15 | Specify any assessment of risk of bias that may affect the cumulative evidence (e.g.. publication bias. selective reporting within studies). | Table 3. page 71 |
| Additional analyses | 16 | Describe methods of additional analyses (e.g.. sensitivity or subgroup analyses. meta-regression). if done. indicating which were pre-specified. | 12 |
| **RESULTS** | | |  |
| Study selection | 17 | Give numbers of studies screened. assessed for eligibility. and included in the review. with reasons for exclusions at each stage. ideally with a flow diagram. | 14 |
| Study characteristics | 18 | For each study. present characteristics for which data were extracted (e.g.. study size. PICOS. follow-up period) and provide the citations. | ESF. Table 2 page |
| Risk of bias within studies | 19 | Present data on risk of bias of each study and. if available. any outcome level assessment (see item 12). | Table 3. page 71 |
| Results of individual studies | 20 | For all outcomes considered (benefits or harms). present. for each study: (a) simple summary data for each intervention group (b) effect estimates and confidence intervals. ideally with a forest plot. | Table 1. page 66 |
| Synthesis of results | 21 | Present results of each meta-analysis done. including confidence intervals and measures of consistency. | Table 2. page 68 |
| Risk of bias across studies | 22 | Present results of any assessment of risk of bias across studies (see Item 15). | Table 3 page 71 |
| Additional analysis | 23 | Give results of additional analyses. if done (e.g.. sensitivity or subgroup analyses. meta-regression [see Item 16]). | Table 2. page 68 |
| **DISCUSSION** | | |  |
| Summary of evidence | 24 | Summarize the main findings including the strength of evidence for each main outcome; consider their relevance to key groups (e.g.. healthcare providers. users. and policy makers). | Page 22-31 |
| Limitations | 25 | Discuss limitations at study and outcome level (e.g.. risk of bias). and at review-level (e.g.. incomplete retrieval of identified research. reporting bias). | Page 31-32 |
| Conclusions | 26 | Provide a general interpretation of the results in the context of other evidence. and implications for future research. | Page 32 |
| **FUNDING** | | |  |
| Funding | 27 | Describe sources of funding for the systematic review and other support (e.g.. supply of data); role of funders for the systematic review. | Page 33 |

**ESF. table 4.** Characteristics of the studies included in the systematic reviews and meta-analysis.

| **NO** | **Authors. years** | **Setting** | **Post COVID period-Months** | **Type of case** | **Type of Control** | **Sample Size** | | | **Age** | | **Specimen** | **Quality score** | **Red point score** | **Findings** |
| --- | --- | --- | --- | --- | --- | --- | --- | --- | --- | --- | --- | --- | --- | --- |
|  |  |  |  |  |  | **Cases M/F** | **Control M/F** | **Total M/F** | **Case-Mean (SD)** | **Control- Mean (SD)** |  |  |  |  |
| 1 | (Holmes, Wist et al. 2021) | Australia | Approximately 3 months | Post-acute phase nonhospitalized COVID-19 patients. | Healthy controls | NA | NA | NA | NA | NA | Plasma | 4 | 14.5 | beta-lipoproteins*, TCHO* |
| 2 | (Silva, Pereira et al. 2023) | Brazil | 70.50 ± 43.10 days post-diagnosis | Mild-to-moderate post-COVID-19 patients | Healthy age-matched controls, tested negative for SARS-CoV-2 | 20 11/9 | 20 14/6 | 40 25/15 | 29.41 (21.90– 34.96) | 29.39 (21.25 – 32.62) | Serum | 5 | 4.5 | beta-lipoproteins*, TCHO* |
| 3 | (Oliván-Blázquez, Bona-Otal et al. 2024) | Spain | 12–24 months | Patients with post-COVID condition | Individuals recovered within three months from acute COVID-19 | 85 17/68 | 85 66/19 | 170 83/87 | 47 (10) | 48 (10) | Plasma | 7 | 12 | HDL*, LDL#, TG#, AIP |
| 4 | (Garrido, Castillo-Peinado et al. 2024) | Spain | 0 to 15 months after infection | Post-COVID condition patients | Asymptomatic individuals | NA | NA | NA | NA | NA | Plasma | 5.5 | 14.5 | HDL*, LDL#, TG#, AIP |
| 5 | (Grote, Schaefer et al. 2024) | Germany | At least 6 months | Patients with post-COVID-19 syndrome or post-vaccination syndrome. | Asymptomatic individuals with serological findings for SARS-CoV-2. | 8 2/6 | 8 0/8 | 16 2/14 | 36 (25–48) | 29 (24–34) | Serum | 2 | 20 | HDL*, LDL#, TG#, AIP |
| 6 | (Agafonova, Elovikova et al. 2024) | Russia | 12 months for the first stage | Patients with a history of COVID-19 confirmed by positive PCR | Subjects with a negative PCR test for COVID-18 | 138 27/111 | 87 18/69 | 225 45/180 | 61 (47-70) | 59 (44-67) | Blood | 7 | 6 | HDL*, LDL#, TG#, AIP |
| 7 | (Alfadda, Rafiullah et al. 2022) | Saudi Arabia | 6 months | With at least one symptom | No symptoms | NA | NA | NA | 51.34 (18,2) | 46.44 (16,8) | Serum | 3.5 | 13.5 | AIP*, HDL*, LDL#, TCHO#, TG* |
| 8 | (Alshehri, AlQahtani et al. 2023) | Saudi Arabia | 1 month | Residual Neurological Deficits | Complete Recovery | 12 9/3 | 13 6/7 | 25 15/10 | NA | NA | NA | 3 | 19 | AIP*, HDL*, LDL#, TCHO#, TG* |
| 9 | (Abdulaziz Alsufyani 2023) | Saudi Arabia | 6‐month | after acute infection | non COVID infected patients | 37 37/0 | 35 35/0 | 72 72/0 | 11 (1202) | 10.86 (1089) | Serum | 7 | 6 | AIP*, HDL*, LDL#, TCHO#, TG* |
| 10 | (Al-Zadjali, Al-Lawati et al. 2024) | Oman | 3-6 months | Long-COVID-19 patients-Mild-Moderate | Healthy individuals who had neither been affected by COVID-19 nor vaccinated. | 88 52/36 | 29 13/16 | 117 65/52 | 39.66 (10.95) | 38.21 (10.55) | Serum | 4 | 14.5 | AIP*, HDL*, LDL#, TCHO#, TG* |
| 11 | (Santana-de Anda, Torres-Ruiz et al. 2024) | Mexico | 7–19 months | Post-acute COVID-19 patients | Without PACS | 19 12/7 | 32 20/12 | 51 32/19 | 48 (29–67) | 49.5 (25–71) | NA | 3 | 19 | AIP*, HDL*, LDL#, TCHO#, TG* |
| 12 | (Aparisi, Martín-Fernández et al. 2022) | Spain | Median follow-up: 514 days. | Hospitalized COVID-19 patients | Healthy individuals admitted for elective surgery. | 108 62/46 | 28 16/12 | 136 78/58 | 68.5 (59-75.5) | 70.5 (NA) | Serum | 4.5 | 11.5 | AIP, HDL#, TCHO#, TG# |
| 13 | (Berezhnoy, Bissinger et al. 2023) | Germany | Median of 152 days, ranging from 47 to 308 days | Long-Term COVID Syndrome | Healthy controls without prior SARS-CoV-2 infection | 33 17/16 | 83 39/44 | 116 56/60 | 56.9 (14.9) | 40.3 (17.4) | Plasma | 4 | 8.5 | AIP, HDL#, TCHO#, TG# |
| 14 | (Chudzik, Lewek et al. 2022) | Poland | 3 months | Patients with Long COVID without comorbidities | Patients without any symptoms after SARS-CoV-2 recovery | 218 67/151 | 270 119/151 | 488 186/302 | 46.03 (11.88) | 44.30 (12.82) | Serum | 3.5 | 7.5 | AIP, HDL#, TCHO#, TG# |
| 15 | (Davico, Martín et al. 2024) | Argentina | 4 to 12 weeks after infection | Patients with post-COVID syndrome | Individuals without a diagnosis of COVID-19 in the last year. | 9 2/7 | 10 3/7 | 19 5/14 | 41 (11) | 31 (10) | Plasma | 5 | 14 | AIP, HDL#, TCHO#, TG# |
| 16 | (Duran, Kurtipek et al. 2022) | Turkey | At least one year after hospitalization. | Patients diagnosed with long COVID | Healthy individuals with a prior history of COVID-19 who fully recovered | 52 26/26 | 80 28/52 | 132 54/78 | 53.6 (12.8) | 49.0 (12.3) | Serum | 2 | 19.5 | ApoB#, ApoB_1#, HDL*, HDL_1*, LDL*, LDL_1#, oxLDL*, oxLDL_1#, TCHO*, TCHO_1# |
| 17 | (Emiroglu, Dicle et al. 2024) | Turkey | 0-11 months | Post-COVID-19 syndrome patients with hyperglycemia or diabetes. | Post-COVID-19 syndrome patients with normal fasting blood glucose. | 103 34/69 | 408 208/200 | 511 242/269 | 57.86 (10.33) | 50.03 (13.24) | Serum | 2.5 | 25 | ApoB#, ApoB_1#, HDL*, HDL_1*, LDL*, LDL_1#, oxLDL*, oxLDL_1#, TCHO*, TCHO_1# |
| 18 | (Erol, Tezcan et al. 2023) | Turkey | At least one year after laboratory-confirmed COVID-19. | Long COVID patients with cardiac symptoms. | Age- and gender-matched individuals without a history of COVID-19. | 105 45/60 | 184 83/101 | 289 128/161 | 56.1 (11.3) | 55.8 (10.7) | Blood | 4 | 6.5 | ApoB#, ApoB_1#, HDL*, HDL_1*, LDL*, LDL_1#, oxLDL*, oxLDL_1#, TCHO*, TCHO_1# |
| 19 | (Gameil, Marzouk et al. 2021) | Egypt | 3-6 months | LongCOVID | HC | 120 67/53 | 120 69/51 | 240 136/104 | 38.29 (5,27) | 37.25 (4,87) | Serum | 5.5 | 9.5 | ApoB#, ApoB_1#, HDL*, HDL_1*, LDL*, LDL_1#, oxLDL*, oxLDL_1#, TCHO*, TCHO_1# |
| 20 | (Kalinskaya, Vorobyeva et al. 2023) | Russia | NA | Post COVID | Controls | 31 8/23 | 27 10/17 | 58 18/40 | 49 (47; 55.5) | 48 (43.5; 54.5) | Serum | 6 | 19.5 | ApoB#, ApoB_1#, HDL*, HDL_1*, LDL*, LDL_1#, oxLDL*, oxLDL_1#, TCHO*, TCHO_1# |
| 21 | (Korkmaz, Çınar et al. 2024) | Turkey | 3 to 6 months | Post-COVID-19 patients without hospitalization | Post-COVID-19 patients without hospitalization | 201 89/112 | 195 104/91 | 396 193/203 | 48.2 (16.3) | 49.2 (14.9) | blood | 4 | 13.5 | ApoB#, ApoB_1#, HDL*, HDL_1*, LDL*, LDL_1#, oxLDL*, oxLDL_1#, TCHO*, TCHO_1# |
| 22 | (Kovarik, Bileck et al. 2023) | Austria | 7 (3-10) | LongCOVID | Healthy Control | 13 4/9 | 13 6/7 | 26 10/16 | 33 (21-53) | 30 (25-43) | Serum | 7.5 | 11.5 | ApoB#, ApoB_1#, HDL*, HDL_1*, LDL*, LDL_1#, oxLDL*, oxLDL_1#, TCHO*, TCHO_1# |
| 23 | (Kuryłowicz, Babicki et al. 2024) | Poland | 12 and 16 weeks after the COVID‐19 | Post-COVID syndrome patients.IST | Post-COVID syndrome (PCS) patients.IST | 69 21/48 | 1280 491/789 | 1349 512/837 | 45.8 (11.6) | 51.6 (13.1) | Serum | 3 | 26 | ApoB#, ApoB_1#, HDL*, HDL_1*, LDL*, LDL_1#, oxLDL*, oxLDL_1#, TCHO*, TCHO_1# |
| 24 | (Labarca, Henríquez-Beltrán et al. 2022) | Chile | 4 months and 1 year | Obstructive Sleep Apnea | Non OSA | 33 22/11 | 23 10/13 | 56 32/24 | 51.4 (11.1) | 38.3 (12.1) | Serum | 4 | 19 | ApoB#, ApoB_1#, HDL*, HDL_1*, LDL*, LDL_1#, oxLDL*, oxLDL_1#, TCHO*, TCHO_1# |
| 25 | (Liu and Kang 2024) | China | 3 months from the onset of symptoms | Post-COVID patients with cardiovascular symptoms | Post-COVID patients without cardiovascular symptoms | 52 27/25 | 152 87/65 | 204 114/90 | 71.5 (64.3, 79.8) | 68.0 (56.3, 80.0) | Blood | 2 | 18 | ApoB#, ApoB_1#, HDL*, HDL_1*, LDL*, LDL_1#, oxLDL*, oxLDL_1#, TCHO*, TCHO_1# |
| 26 | (Mora, Kogut et al. 2023) | United States | Symptoms exceeding 28 days were classified as long COVID | Long COVID | No COVID | 94 28/66 | 104 49/55 | 198 77/121 | 43.8 (9.9) | 44.8 (12.3) | Blood | 5 | 21 | TCHO* |
| 27 | (Mouchati, Durieux et al. 2023) | United States | Median follow-up was 292 days (IQR: 172, 518) for COVID+ No PASC and 229 days (IQR: 147, 478) for COVID+ PASC+ | Post-COVID patients with or without PASC symptoms. | COVID-negative participants | 85 35/50 | 258 157/101 | 343 192/151 | 47.81 (13.49) | 43.68 (13.69) | Plasma | 5 | 18.5 | AIP#, AIP_1#, HDL*, HDL_1*, LDL#, LDL_1#, TCHO#, TCHO_1#, TG#, TG_1# |
| 28 | (Oikonomou, Lampsas et al. 2023) | Greece | 6 months | Convalescent COVID-19 patients. | Age-, sex-, and cardiovascular risk factor-matched non-COVID-19 individuals | 34 26/8 | 34 23/11 | 68 49/19 | 57.2 (12.9) | 57.4 (12.8) | Serum | 5 | 9.5 | AIP#, AIP_1#, HDL*, HDL_1*, LDL#, LDL_1#, TCHO#, TCHO_1#, TG#, TG_1# |
| 29 | (de Oliveira, de Ávila et al. 2022) | Brazil | Median of 138 days (IQR 90–201 days). | Long COVIDN | No long COVIDN | 369 180/189 | 70 41/29 | 439 221/218 | 57 (46-66) | 62 (49-71) | NA | 3 | 14.5 | AIP#, AIP_1#, HDL*, HDL_1*, LDL#, LDL_1#, TCHO#, TCHO_1#, TG#, TG_1# |
| 30 | (Орлова, Ломайчиков et al. 2021) | Russia | NA | Patients with acute coronary syndrome who had previously suffered from COVID-19 | Patients with ACS without a history of COVID-19 | 109 109/0 | 76 76/0 | 185 185/0 | 64.4 (62, 0; 66, 9) | 68.2 (66, 2; 71, 4) | Serum | 2 | 20 | AIP#, AIP_1#, HDL*, HDL_1*, LDL#, LDL_1#, TCHO#, TCHO_1#, TG#, TG_1# |
| 31 | (Paris, Palomba et al. 2023) | Italy | Within 2 months | Post-COVID-19 patients with long-COVID conditions | Age- and sex-matched healthy volunteers | 38 35/3 | 38 35/3 | 76 70/6 | 58.82 (10.08) | 57.93 (11.23) | NA | 4 | 17 | AIP#, AIP_1#, HDL*, HDL_1*, LDL#, LDL_1#, TCHO#, TCHO_1#, TG#, TG_1# |
| 32 | (Paul, Paul et al. 2023) | India | More than 6 months | Post-COVID-19 patients | Age- and sex-matched nonsmokers | 108 57/51 | 108 52/56 | 216 109/107 | NA | NA | Blood | 4 | 12.5 | AIP#, AIP_1#, HDL*, HDL_1*, LDL#, LDL_1#, TCHO#, TCHO_1#, TG#, TG_1# |
| 33 | (Zerón-Rugerio, Zaragozá et al. 2024) | Spain | NA | Patients with post-COVID ME-CFS | Matched healthy controls | 23 10/13 | 31 10/21 | 54 20/34 | 49.61 (2.09) | 43.06 (1.99) | Plasma | 4.5 | 9 | AIP#, AIP_1#, HDL*, HDL_1*, LDL#, LDL_1#, TCHO#, TCHO_1#, TG#, TG_1# |
| 34 | (Stepanova, Driianska et al. 2024) | Ukraine | One year after COVID-19 infection | Long COVID comorbid with Hemodialysis | Hemodialysis patients who had fully recovered from COVID-19 | 45 28/17 | 35 17/18 | 80 45/35 | 56 (44–62) | 55 (45–64) | Serum | 6 | 9 | AIP#, AIP_1#, HDL*, HDL_1*, LDL#, LDL_1#, TCHO#, TCHO_1#, TG#, TG_1# |
| 35 | (Sumbalová, Kucharská et al. 2022) | Slovakia | NA | Post COVID | Healthy Control | 14 8/6 | 15 6/9 | 29 14/15 | 51.3 (2.3) | 51.3 (2.3) | Plasma | 4.5 | 14 | AIP#, AIP_1#, HDL*, HDL_1*, LDL#, LDL_1#, TCHO#, TCHO_1#, TG#, TG_1# |
| 36 | (Szczerbiński, Okruszko et al. 2023) | Poland | Approximately six months | Adults with a history of SARS-CoV-2 infection confirmed by PCR and hospitalized during the acute phase. | Age ,sex and BMI-matched participants from a population study conducted pre-pandemic. | 39 13/26 | 39 13/26 | 78 26/52 | 48.64 (2.24) | 48.79 (2.22) | Serum | 6 | 9 | AIP#, AIP_1#, HDL*, HDL_1*, LDL#, LDL_1#, TCHO#, TCHO_1#, TG#, TG_1# |
| 37 | (Tong, Yan et al. 2022) | China | 1 year (375.0 ± 11.0 days) | COVID-19 survivors one year after discharge | Age- and gender-matched healthy medical staff | 54 19/35 | 119 46/73 | 173 65/108 | 54 (41-61) | 52 (42-61) | Serum | 6 | 12 | AIP#, ApoA1*, ApoA2*, ApoB#, FCHO*, HDL#, LDL*, TG#, VLDL# |
| 38 | (Tudoran, Bende et al. 2023) | Romania | Median time elapsed since COVID-19 diagnosis was 56 days for group A and 63 days for group B. | Group A: 67 Women with MS and a History of COVID-19 | Healthy, premenopausal, age-matched women who never had COVID-19. | 67 0/67 | 40 0/40 | 107 0/107 | 50.59 (4.53) | 49.47 (5.14) | Serum | 4.5 | 14.5 | AIP#, ApoA1*, ApoA2*, ApoB#, FCHO*, HDL#, LDL*, TG#, VLDL# |
| 39 | (Verma, Ramayya et al. 2022) | United States | Mean 332 ± 130 days | Patients with post-COVID-19 syndrome PASC-CVS | Healthy controls | 23 2/21 | 23 10/13 | 46 12/34 | 46 (11) | 56 (12) | Serum | 4 | 6.5 | AIP#, ApoA1*, ApoA2*, ApoB#, FCHO*, HDL#, LDL*, TG#, VLDL# |
| 40 | (Vollrath, Matits et al. 2023) | Germany | 3.8 ± 2.68 months | Previously SARS-CoV-2-infected athletes | Healthy athletic | 59 30/29 | 31 12/19 | 90 42/48 | 34.5 (12.2) | 31.9 (10.4) | Blood | 5 | 6.5 | AIP#, ApoA1*, ApoA2*, ApoB#, FCHO*, HDL#, LDL*, TG#, VLDL# |
| 41 | (Vyas, Joshi et al. 2023) | India | 1 year | Post COVID-Hypertensive | postCOVID-Normotensive | 80 60/20 | 168 109/59 | 248 169/79 | 52.39 (12.64) | 50.35 (13.91) | Blood | 4 | 16.5 | AIP#, ApoA1*, ApoA2*, ApoB#, FCHO*, HDL#, LDL*, TG#, VLDL# |
| 42 | (Xuereb, Borg et al. 2023) | Malta | Approximately 6 months (median follow-up: 173.5 days). | Patients previously diagnosed with COVID-19 infection. | Age- and gender-matched individuals who tested negative for COVID-19. | 174 69/105 | 75 34/41 | 249 103/146 | 45.5 (35-58.75) | 44 (37.5-56.5) | Serum | 5.5 | 12.5 | AIP#, ApoA1*, ApoA2*, ApoB#, FCHO*, HDL#, LDL*, TG#, VLDL# |
| 43 | (Yamamoto, Otsuka et al. 2023) | Japan | at least 6 months | Long COVID with ME-CSF | No fatigue | 50 24/26 | 95 38/57 | 145 62/83 | 42 (30.3–51.8) | 43 (29.5-51) | Blood | 4 | 18 | AIP#, ApoA1*, ApoA2*, ApoB#, FCHO*, HDL#, LDL*, TG#, VLDL# |
| 44 | (Zisis, Durieux et al. 2023) | United States | Average 323–364 days | Post-acute COVID-19 patients (with and without PASC symptoms) | COVID-negative individuals | 68 45/23 | 37 25/12 | 105 70/35 | 43.23 (14.33) | 43.41 (14.04) | Blood | 7 | 8 | AIP#, ApoA1*, ApoA2*, ApoB#, FCHO*, HDL#, LDL*, TG#, VLDL# |

*: Indicates that patients have reduced level of the measured metabolite compared to healthy control (negative SMD)

^#:^ Indicates that patients have increased level of the measured metabolites compared to healthy control (positive SMD)

TG: Triglyceride, HDL: High-density lipoprotein, LDL: Low-density lipoprotein, VLDL: Very low-density lipoprotein, Apo: Apolipoprotein, TCHO: Total cholesterol, AIP: atherogenic index of plasma.

**ESF. Table 5**. Results of Meta-regression

| Variables | No. of Studies | Covariates | 1-sided p-value | Z-Value |
| --- | --- | --- | --- | --- |
| TG/HDL | 33 | Male-Case | 0.037 | -1.78 |
| CRI-1 | 28 | Less than 3 Months | 0.000 | 3.99 |
|  | 9 | SBP | 0.010 | 2.31 |
| (TG+LDL+VLDL)/(HDL+ApoA) | 35 | Male-Cases | 0.029 | -1.89 |
|  | 37 | Plasma | 0.049 | 1.65 |
|  | 9 | Heart Disease | 0.016 | 2.13 |
| HDL | 26 | Less than 3 Months | 0.007 | -2.44 |
| TCHO | 30 | Less than 3 Months | 0.012 | 2.25 |
|  | 4 | Hospitalization during acute stage of illness | 0.008 | 2.38 |
| TG | 28 | More than 6 Months | 0.014 | -2.18 |
|  | 31 | Male-Cases | 0.010 | -2.33 |
|  | 33 | Plasma | 0.031 | 1.85 |
| LDL | 31 | Latitude | 0.016 | -2.14 |
|  | 5 | Medication Free | 0.006 | 2.51 |

CRI: Castelli risk index, TG: Triglyceride, HDL: High-density lipoprotein, LDL: Low-density lipoprotein, VLDL: Very low-density lipoprotein, Apo: Apolipoprotein, TCHO: Total cholesterol, SBP: systolic blood pressure.

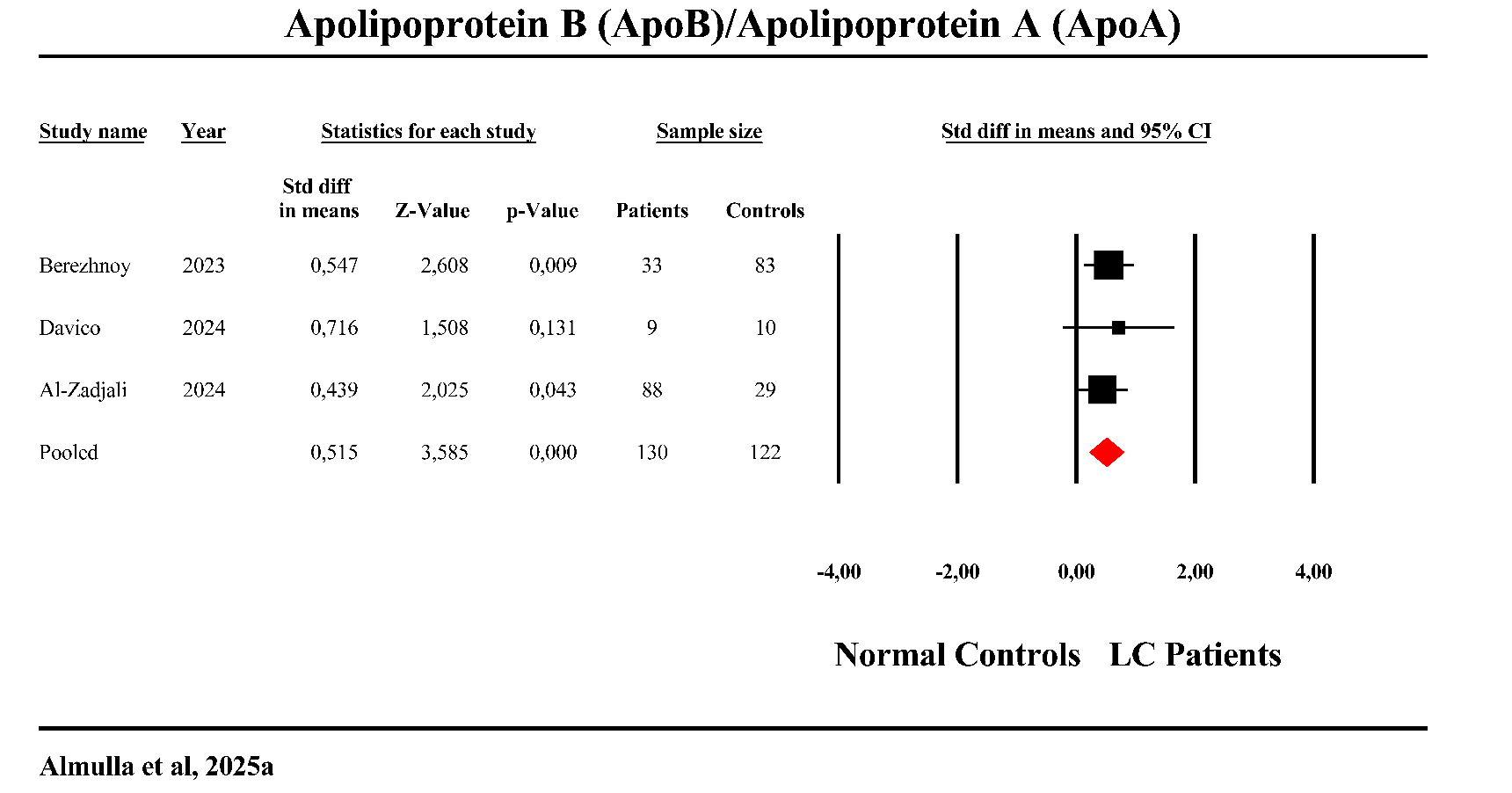

**ESF. Figure 1**. Forest plot of apolipoprotein B (ApoB)/apolipoprotein A (ApoA) in the patients with Long COVID (LC) versus normal controls.

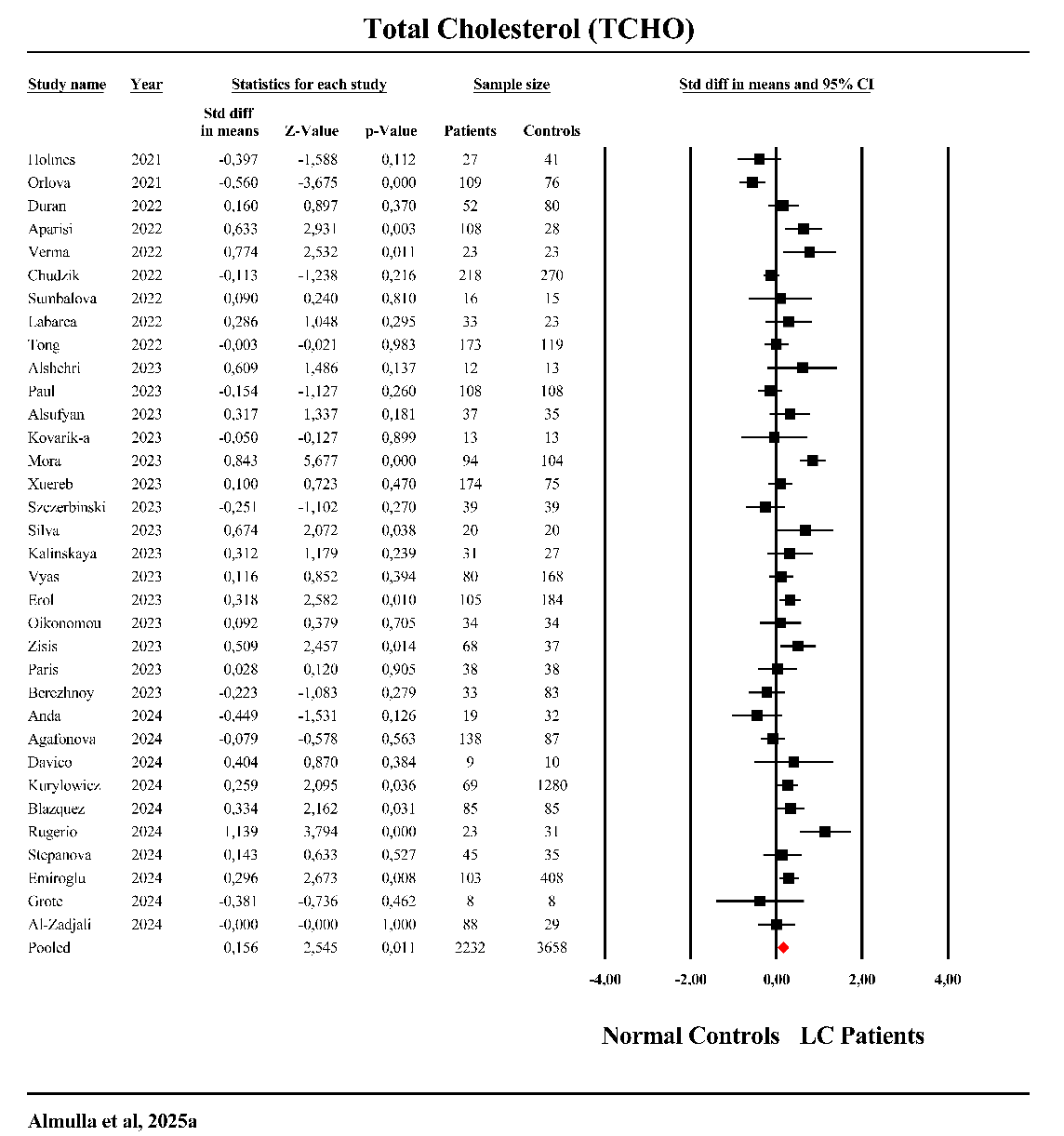

**ESF. Figure 2**. Forest plot of total cholesterol (TCHO) in the patients with Long COVID (LC) versus normal controls.

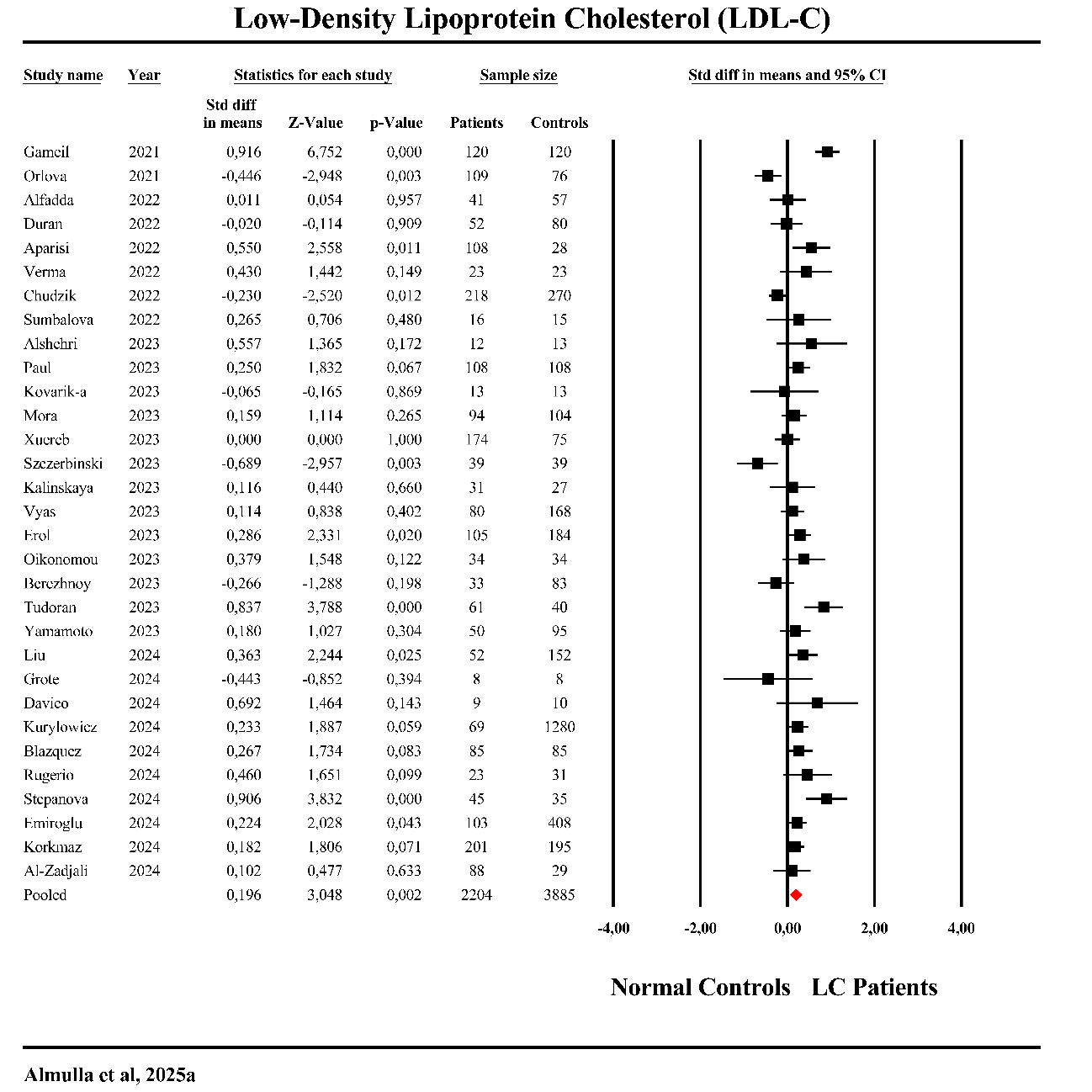

**ESF. Figure 3**. Forest plot of low-density lipoprotein (LDL) in the patients with Long COVID (LC) versus normal controls.

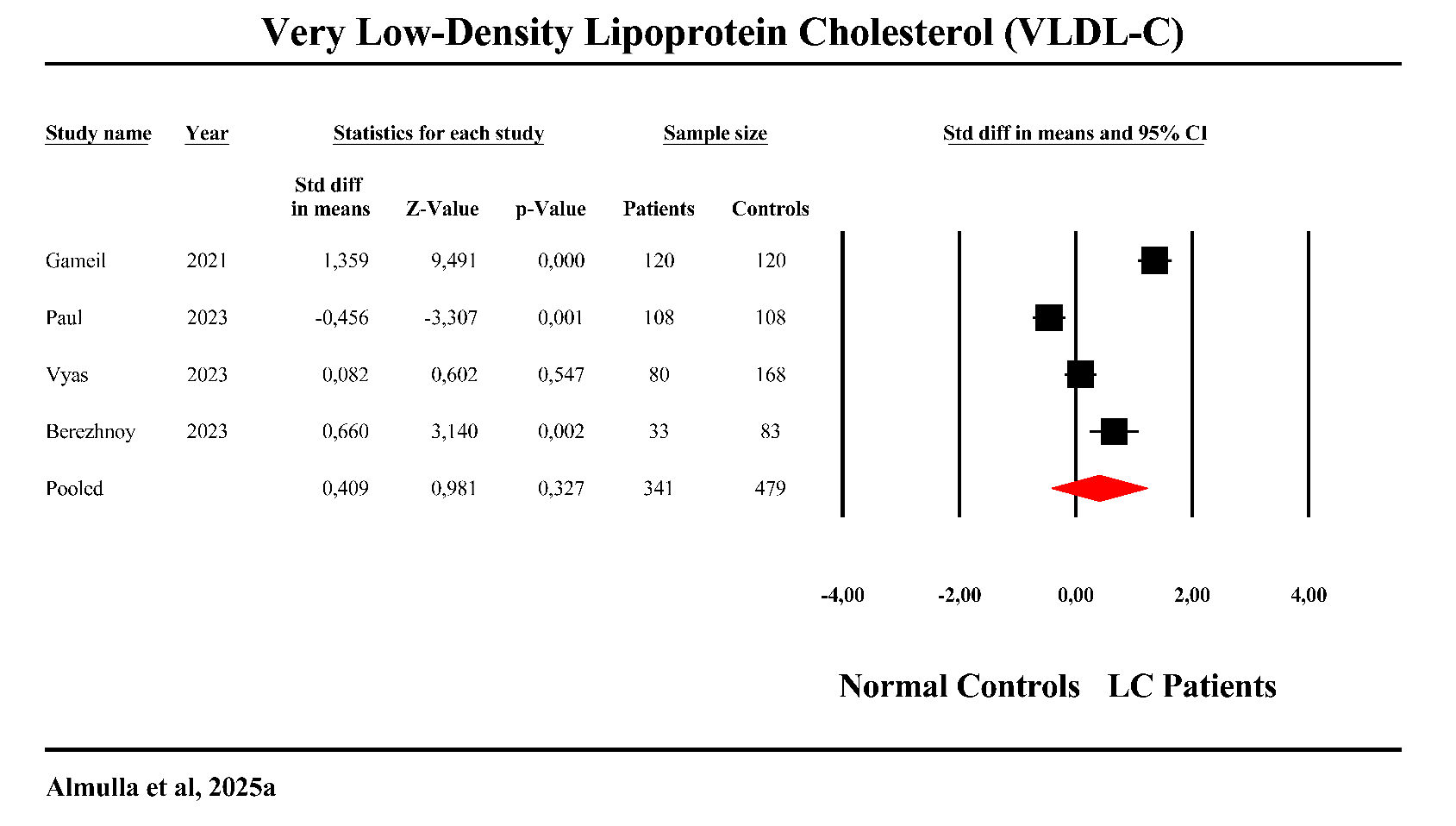

**ESF. Figure 4**. Forest plot of very low-density lipoprotein (VLDL) in patients with Long COVID (LC) versus normal control.

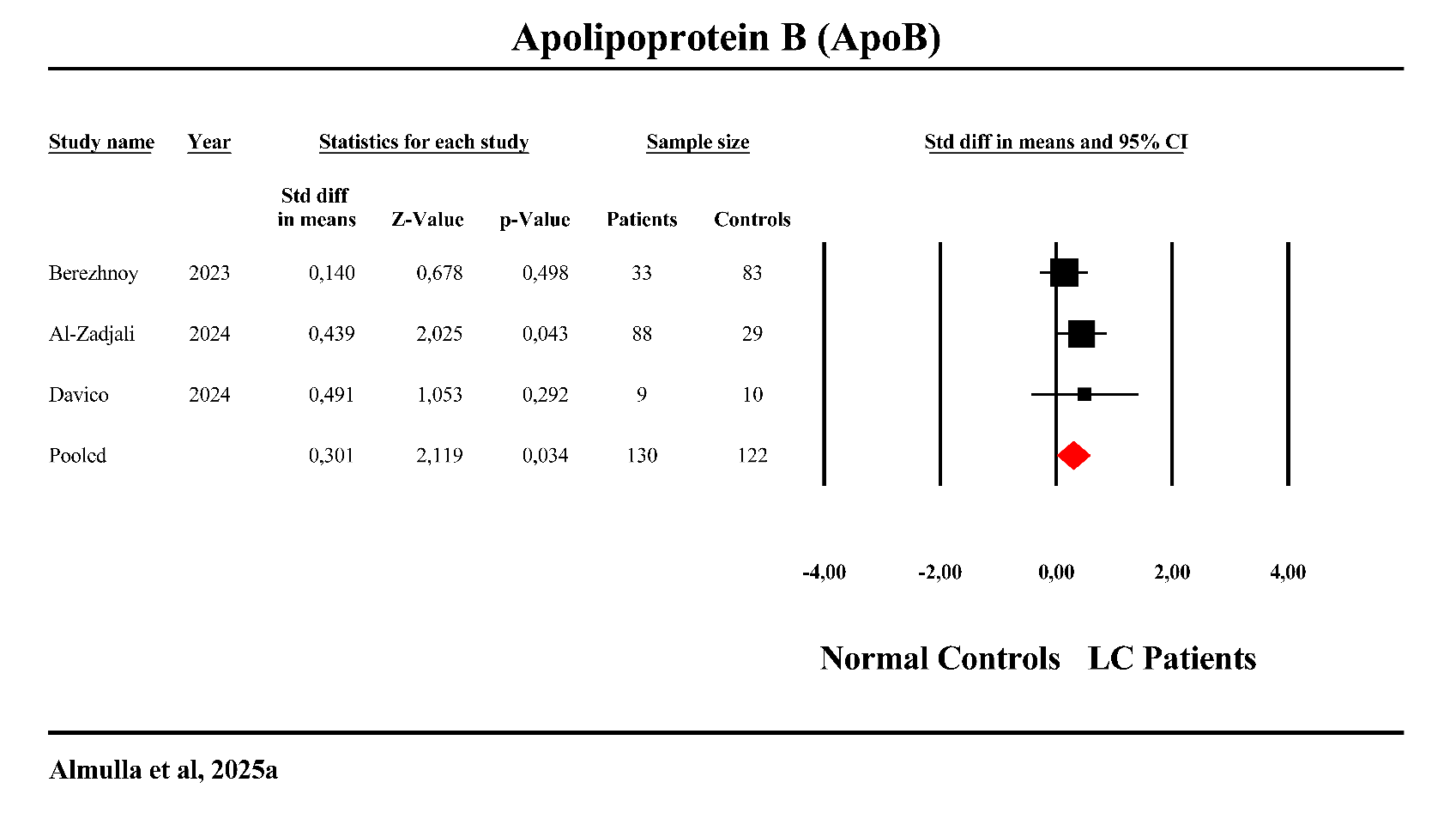

**ESF. Figure 5**. Forest plot of apolipoprotein B (ApoB) in patients with Long COVID (LC) versus normal control.

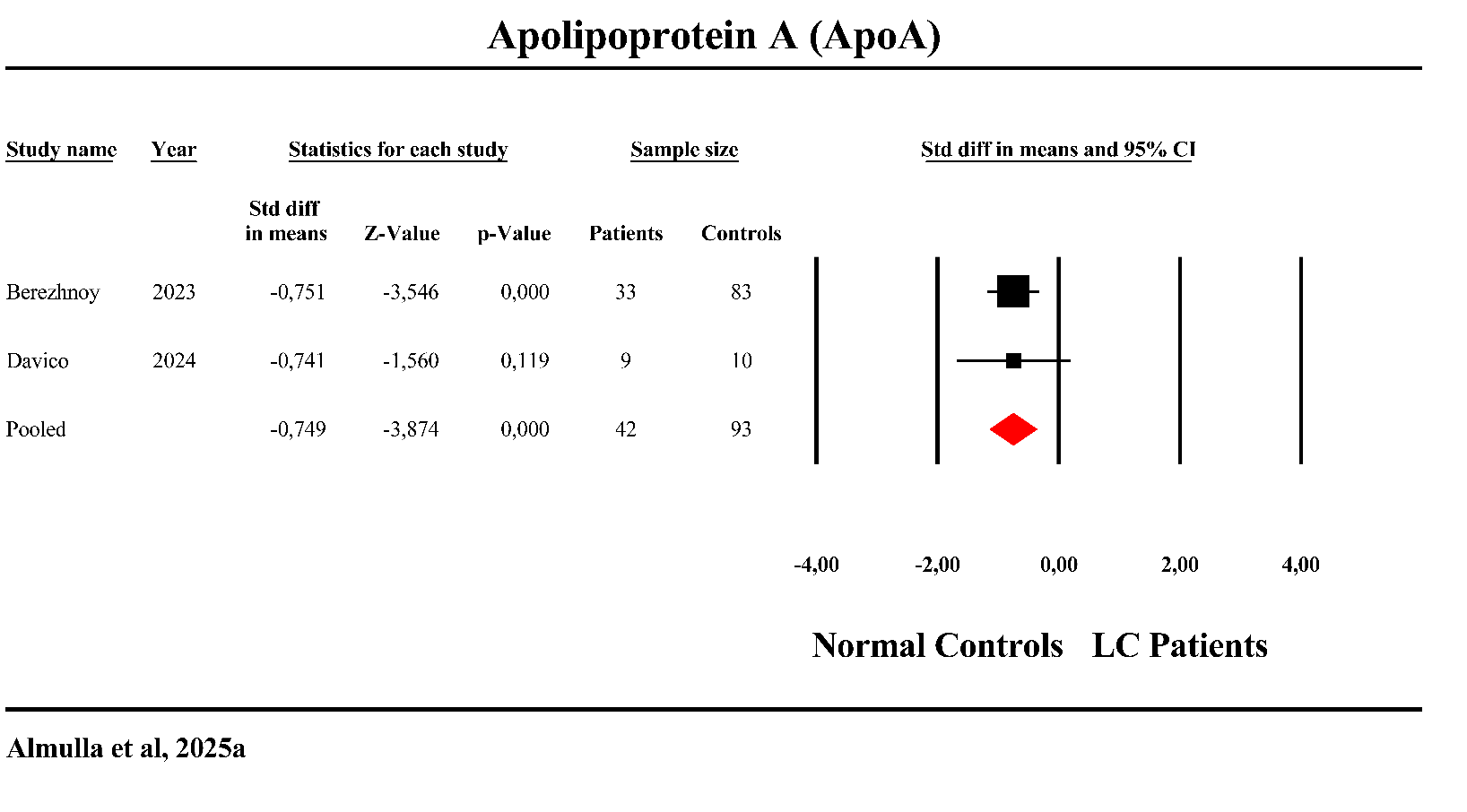

**ESF. Figure 6**. Forest plot of apolipoprotein A (ApoA) in patients with Long COVID (LC) versus normal control.
